# Genome-wide association study of REM sleep behavior disorder identifies novel loci with distinct polygenic and brain expression effects

**DOI:** 10.1101/2021.09.08.21254232

**Authors:** Lynne Krohn, Karl Heilbron, Cornelis Blauwendraat, Regina H. Reynolds, Eric Yu, Konstantin Senkevich, Uladzislau Rudakou, Mehrdad A. Estiar, Emil K. Gustavsson, Kajsa Brolin, Jennifer A. Ruskey, Kathryn Freeman, Farnaz Asayesh, Ruth Chia, Isabelle Arnulf, Michele T.M. Hu, Jacques Y. Montplaisir, Jean-François Gagnon, Alex Desautels, Yves Dauvilliers, Gian Luigi Gigli, Mariarosaria Valente, Francesco Janes, Andrea Bernardini, Birgit Högl, Ambra Stefani, Abubaker Ibrahim, Karel Sonka, David Kemlink, Wolfgang Oertel, Annette Janzen, Giuseppe Plazzi, Francesco Biscarini, Elena Antelmi, Michela Figorilli, Monica Puligheddu, Brit Mollenhauer, Claudia Trenkwalder, Friederike Sixel-Döring, Valérie Cochen De Cock, Christelle Charley Monaca, Anna Heidbreder, Luigi Ferini-Strambi, Femke Dijkstra, Mineke Viaene, Beatriz Abril, Bradley F. Boeve, 23andMe Research Team, Sonja W. Scholz, Mina Ryten, Sara Bandres-Ciga, Alastair Noyce, Paul Cannon, Lasse Pihlstrøm, Mike A. Nalls, Andrew B. Singleton, Guy A. Rouleau, Ronald B. Postuma, Ziv Gan-Or

## Abstract

Rapid eye movement (REM) sleep behavior disorder (RBD), enactment of dreams during REM sleep, is an early clinical symptom of alpha-synucleinopathies. RBD also defines more severe forms of alpha-synucleinopathies. The genetic background of RBD and its underlying mechanisms are not well understood. Here, we performed the first genome-wide association study of RBD, identifying five RBD risk loci. Expression analyses highlight *SNCA-AS1* and *SCARB2* differential expression in different brain regions in RBD, with *SNCA-AS1* further supported by colocalization analyses. Genetic risk score and other analyses provide further insights into RBD genetics, highlighting RBD as a unique subpopulation that will allow future early intervention.

## INTRODUCTION

Rapid eye movement (REM) sleep behavior disorder (RBD), defined as loss of muscle atonia and dream enactment during REM sleep, is one of the most unique conditions in neurology^1^. Isolated RBD (iRBD), defined as having RBD without other significant clinical neurological signs, is the only early highly predictive clinical marker for some neurodegenerative diseases. Over 80% of iRBD patients will convert to overt neurodegeneration within 10-15 years on average, most commonly to Parkinson’s disease (PD) or dementia with Lewy bodies (DLB), or in rare cases, multiple system atrophy (MSA) or another neurodegenerative disorder.^2,3^ Although they demonstrate phenotypic differences, PD, DLB, and MSA are all alpha-synucleinopathies: disorders characterized by accumulation of the protein alpha-synuclein in the brain. Therefore, iRBD is considered a prodromal alpha-synucleinopathy which offers a unique opportunity to identify these conditions at a much earlier stage.^4^

There is strong evidence that iRBD also represents distinct, more severe subtypes of alpha-synucleinopathies. Approximately 30-60% of PD patients have RBD, including both iRBD and RBD as a symptom occurring after PD diagnosis (symptomatic RBD, sRBD).^3^ In this manuscript, we will use “RBD” to refer to all instances of RBD regardless of at which stage symptoms present, and iRBD or sRBD to specify before or after overt neurodegeneration diagnosis, respectively. The presence of RBD is currently the strongest predictor for the development of dementia in PD^5^ and is associated with more rapid progression of non-motor symptoms.^3^ RBD is more frequent in DLB, found in approximately 50-80% of all cases. While the phenotypic differences in DLB with or without RBD are less pronounced, those diagnosed with iRBD before developing DLB tend to have increased severity of DLB symptoms and more rapid deterioration.^6^ MSA patients also have a high prevalence of RBD, estimated at 75-95%, 40% of which have iRBD prior to the onset of MSA symptoms. Those with iRBD preceding MSA symptoms may have more frequent autonomic onset of MSA, less frequent parkinsonism at MSA onset, and a more severe disease course.^7^ Overall, RBD, and specifically iRBD, appears to represent a more malignant subtype of alpha-synucleinopathies.

Thus far, the genetics of RBD has only been studied through the candidate gene approach. To better understand RBD and early alpha-synucleinopathy genetics and potential mechanisms, we performed a genome-wide association study (GWAS) on 2,843 cases and 139,636 controls. We further examined the biological implications of the nominated risk loci through pathway analysis, investigated variant effects on gene expression, and assessed the cumulative risk using polygenic risk score (PRS). Using the GWAS summary statistics, we studied the genetic relationship between RBD and the synucleinopathies to which it progresses, as well as conditions and exposures of interest using linkage disequilibrium (LD) score regression and Mendelian randomization (MR).

## RESULTS

### Genome-wide association study identifies five RBD loci

To identify genetic risk loci across the genome associated with RBD, we performed a case-control GWAS of iRBD (N cases=1,061, N controls=8,386) and a case-control GWAS from 23andMe, Inc. using PD patients with probable RBD (pRBD) and controls (PD+pRBD, diagnosed by a validated questionnaire, N cases=1,782, N controls=131,250), meta-analyzed for a total of 2,843 cases and 139,636 controls. After filtration, imputation, and meta-analysis, a total of 11,536,573 variants were examined. We tested for systemic biases using the genomic inflation factor (lambda) and LD-score regression, with satisfactory results (lambda=1.06, lambda1000=1.01, LD intercept= -0.01). With LD-score regression, the liability-scale narrow-sense heritability of iRBD based on common variants is calculated at 12.3% (standard error=0.07), similar to the recently reported 10.8% heritability for DLB.^8^

We identified novel RBD-associated loci near *SCARB2* and *INPP5F*, and replicated known RBD associations near *SNCA*,^9^ *GBA*,^10,11^ and *TMEM175*^*12*^ (Table 1, Figure 1). Conditional-joint analysis identified a secondary associated single nucleotide polymorphism (SNP) in *GBA*, confirming RBD risk association with known PD nonsynonymous variants p.Glu326Lys and p.Asn370Ser. These five loci have also been implicated in PD,^13^ however the RBD-associated SNPs in *SNCA* and *SCARB2* are not in LD with the top PD-associated SNPs in these loci, and are thus considered independent. Additionally, SNPs associated with PD or DLB-associated SNPs in notable GWAS loci, such as *MAPT, LRRK2*, and *APOE*^8^ are not associated with RBD despite sufficient power to detect them, suggesting that RBD has a distinct and only partially overlapping genetic background with PD and DLB.

**TABLE 1.** Independent RBD risk loci nominated by GWAS meta-analysis. *SNP: single nucleotide polymorphism; Eff: effect; Ref: reference; MAF: minor allele frequency; SE: standard error; Het: heterogeneity*. We identified 6 independent variants significantly associated with RBD according to the standard GWAS multiple testing correction (p<5E-08). The high heterogeneity found in the SNCA locus could be attributed to the stronger effect size in iRBD (beta=0.40) compared to PD+pRBD (beta=0.15).

| Position (hg19) | SNP | Closest gene | Eff Allele | Ref Allele | MAF | Beta | SE | P-value | Het I<sup>2</sup> (%) |
| --- | --- | --- | --- | --- | --- | --- | --- | --- | --- |
| 4:90757272 | rs3756059 | *SNCA* | A | G | 0.50 | 0.23 | 0.03 | 3.02E-16 | 94.4 |
| 1:155205378 | rs12752133 | *GBA* | T | C | 0.01 | 0.74 | 0.10 | 4.87E-14 | 0 |
| 1:155205634 | rs76763715 | *GBA* | T | C | 0.99 | -1.04 | 0.16 | 1.68E-10 | 0 |
| 4:951947 | rs34311866 | *TMEM175* | T | C | 0.81 | -0.20 | 0.04 | 4.41E-09 | 0 |
| 10:121536327 | rs117896735 | *INPP5F* | A | G | 0.01 | 0.59 | 0.10 | 4.70E-09 | 0.60 |
| 4:77132634 | rs7697073 | *SCARB2* | T | C | 0.34 | 0.16 | 0.03 | 2.21E-08 | 0 |

**FIGURE 1.**
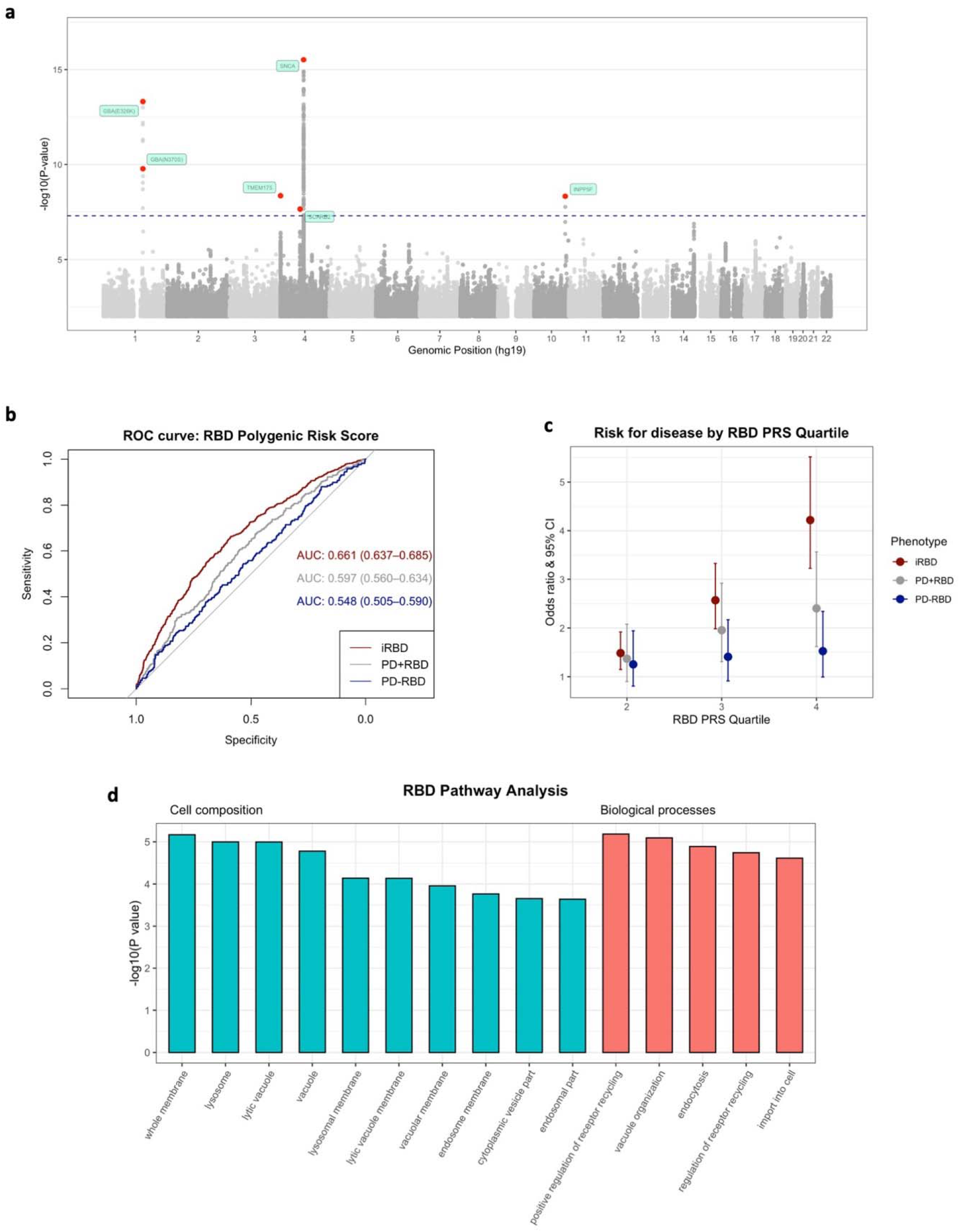
Summary of GWAS findings in the RBD meta-analysis. Manhattan plot. **(a)** The Manhattan plot highlights the 6 GWAS-nominated loci after meta-analysis. Each point represents the log adjusted p-value at each genomic site. The points in red show the top variant at that particular locus, as well as any secondary associations identified by COJO. The GWAS-significant p-value threshold of 5E-08 is visualized with the dashed line. Polygenic risk scores for RBD were calculated using FDR-corrected GWAS variants in 3 cohorts: idiopathic RBD, PD+pRBD, and PD-pRBD, each with controls. **Predictive power of RBD polygenic risk score. (b)** The predictive power of the PRS in each cohort was assessed with area under the curve (AUC) and 95% confidence intervals. **(c)** The PRS for each cohort were divided in quartiles and analyzed against case status with logistic regression. The odds ratios and 95% confidence intervals are visualized here as compared to the lowest quartile (lowest 25% of scores). **(d) Pathways associated with genes nominated by RBD meta-analysis**. Bars represent the unadjusted p-values for pathways nominated by functional enrichment analysis in cellular component (blue) and biological processes (red). All pictured pathways are significant after FDR multiple testing correction.

### Differential gene expression in different brain regions may drive the independent associations of SNCA and SCARB2 in RBD and PD

We found that although the *SNCA* and *SCARB2* loci are important in both RBD and PD, the associations with both conditions in these loci are driven by independent variants. In the *SNCA* locus, rs3756059 (in the 5’ region of the gene) was associated with RBD in the current GWAS and with DLB.^8^ In contrast, rs356182 (in the 3’ region), which is not in LD with rs3756059 (R^2^=0.17, D’=0.56), is the most significant of all association signals in GWAS of PD risk,^13^ yet showed no association with iRBD. Similarly, in the *SCARB2* locus, rs7697073 is associated with RBD and rs6825004 is associated with PD, and the two SNPs are not in LD (R^2^=0.06, D’=0.26). We therefore hypothesized that the different SNPs in these loci associated with RBD and PD may be associated with differential expression patterns of their respective genes in different brain regions. To examine this hypothesis, we used the publicly available Genotype-Tissue Expression (GTEx) consortium v8 online portal (https://www.gtexportal.org/home/)^14^ to investigate how each variant affects gene expression in specific brain tissues. We examined the effects of these variants on the expression of *SCARB2, SNCA* and *SNCA* antisense-1 (*SNCA-AS1*), since variants in *SNCA* have previously been linked to *SNCA-AS1* expression.^8^ The expression quantitative trait loci (eQTL) effect size (ES) reported here is the slope of the linear regression of normalized expression data versus the genotype status using single-tissue eQTL analysis, performed by the GTEx consortium. Comparative results are visualized in Figure 2.

**FIGURE 2.**
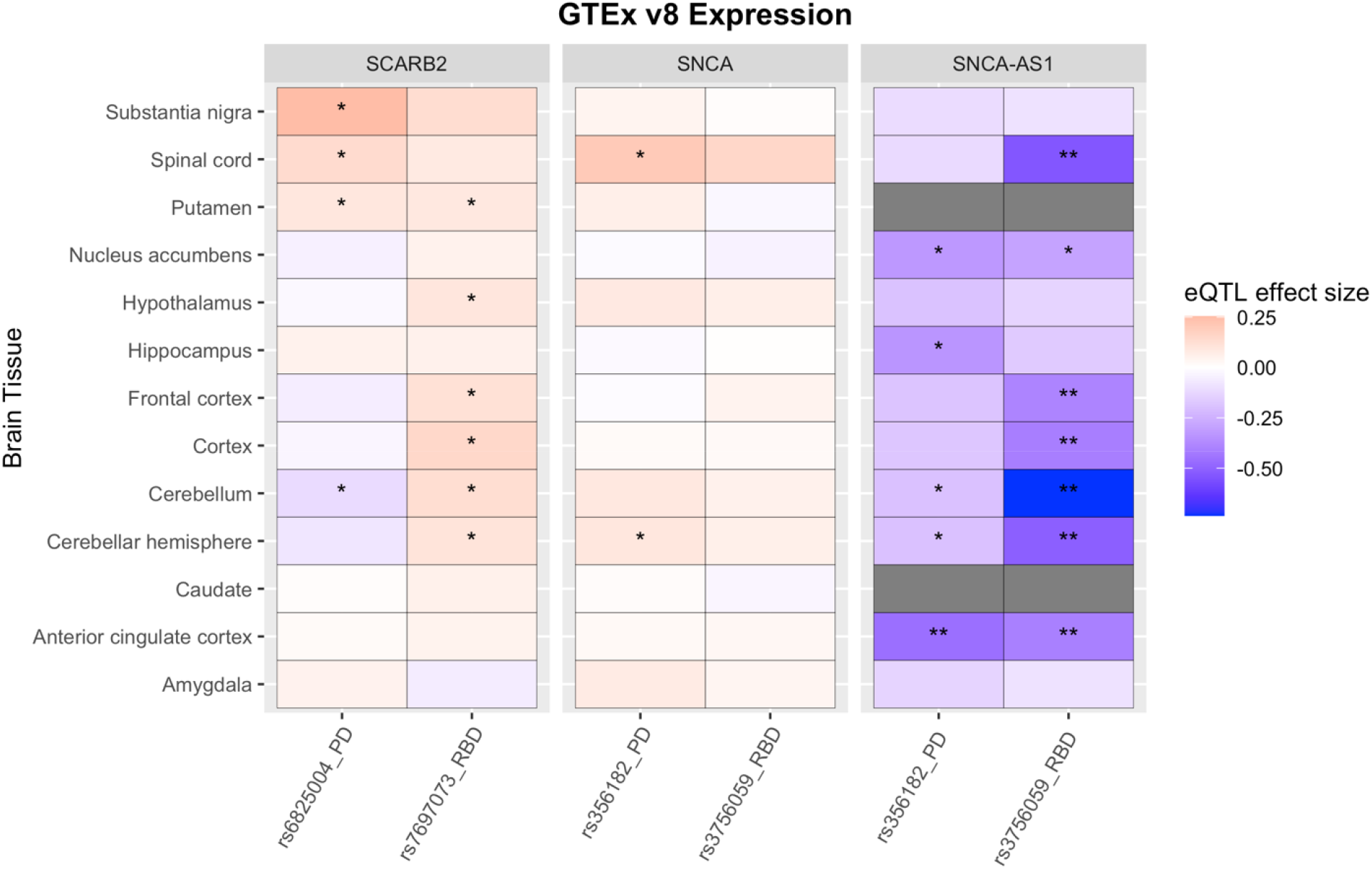
Expression data from GTEx version 8 for RBD and PD top variants in differing loci. All data was extracted from the GTEx online portal (https://www.gtexportal.org/). Nominal associations are indicated with * (p<0.05) while Bonferroni-corrected significant associations are indicated with **. Dark grey indicates missing data in these tissues.

In the *SNCA/SNCA-AS1* locus, the RBD risk variant rs3756059 is most strongly associated with increased expression for *SNCA-AS1* expression in multiple cortical regions (frontal cortex ES=-0.39, p=6.3E-05; anterior cingulate cortex ES=-0.41, p=2.6E-04), the cerebellum (ES=-0.74, p=9.9E-19) and the spinal cord (ES=-0.54, p=2.0E-05), all with statistical significance after correction for multiple comparisons. The PD variant rs356182 is only associated with *SNCA-AS1* expression in the anterior cingulate cortex (ES=-0.46, p=2.5E-05). The RBD risk variant at the *SCARB2* locus (rs7697073) is most strongly associated with increased expression in the cortex (ES=0.15, p=1.4E-03), while the *SCARB2* PD risk variant (rs6825004) is most strongly associated with substantia nigra increased expression (ES=0.25, p=4.6E-04). The PD variant is also associated with decreased *SCARB2* expression in the cerebellum, while the opposite is shown for the RBD variant. Of note, *SCARB2* associations are nominal and lack significance after GTEx v8 multiple testing correction.

### Colocalization analyses demonstrate tissue and cell-specific differential effects of RBD-associated variants

We further investigated RBD loci effects on gene expression using colocalization analysis to determine whether association signals for RBD risk and regulation of gene expression in the whole brain or blood are driven by a shared causal variant. eQTLs were obtained from PsychENCODE^15^ and eQTLGen,^16^ large human brain and blood eQTL datasets, respectively. In brain, we found one RBD locus with strong evidence of colocalization with eQTLs regulating *SNCA-AS1* expression (posterior probability of hypothesis 4 [one shared variant between RBD GWAS and eQTL, PPH4] = 0.89; Figure 3A; Supplementary Table 1). Using beta coefficients derived from the RBD GWAS and PsychENCODE eQTLs, we found that SNPs in the region surrounding *SNCA-AS1* tended to show an inverse relationship between RBD risk and *SNCA-AS1* expression, suggesting that reduced RBD risk is associated with increased *SNCA-AS1* expression (Supplementary Fig. 4), which in turn may be associated with reduced alpha-synuclein protein level.

**FIGURE 3.**
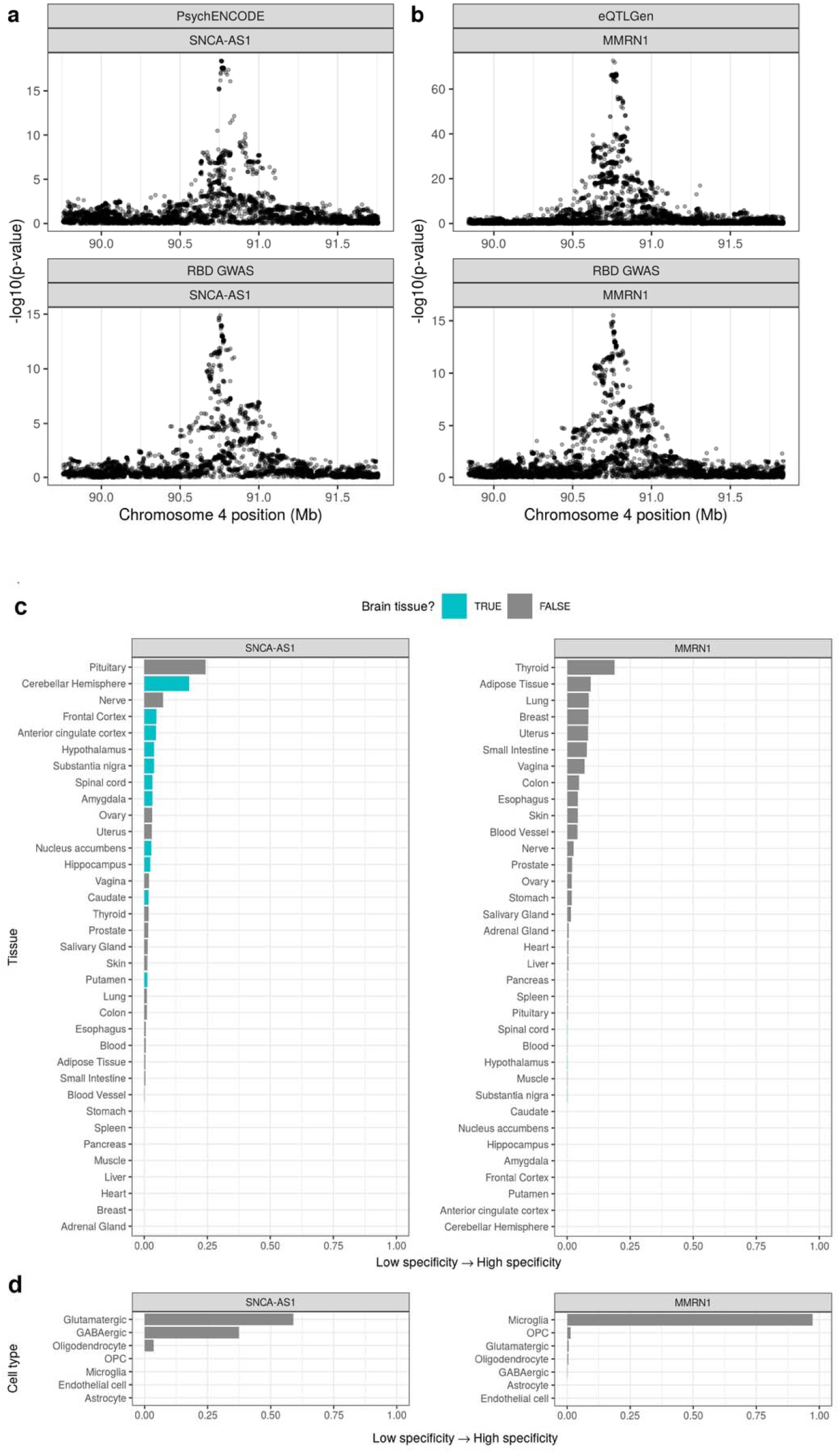
Regional association plots for eQTL and RBD GWAS colocalizations and tissue and cell-type specificity of *MMRN1* and *SNCA-AS1*. Regional association plots for eQTL (upper pane) and RBD GWAS association signals (lower pane) in the regions surrounding **(a)** *SNCA-AS1* (PPH4 = 0.89) and **(b)** *MMRN1* (PPH4 = 0.86). eQTLs are derived from **(a)** PsychENCODE’s analysis of adult brain tissue from 1387 individuals or **(b)** the eQTLGen meta-analysis of 31,684 blood samples from 37 cohorts. In **(a)** and **(b)**, the x-axis denotes chromosomal position in hg19, and the y-axis indicates association p-values on a -log_10_ scale. Plot of *SNCA-AS1* and *MMRN1* specificity in **(c)** 35 human tissues (GTEx dataset) and **(d)** 7 broad categories of cell type derived from human middle temporal gyrus (AIBS dataset). Specificity represents the proportion of a gene’s total expression attributable to one cell type/tissue, with a value of 0 meaning a gene is not expressed in that cell type/tissue and a value of 1 meaning that a gene is only expressed in that cell type/tissue. In **(c)** tissues are colored by whether they belong to the brain. In **(c)** and **(d)**, tissues and cell types have been ordered by specificity from high to low.

In blood, we found evidence of colocalizations between the same RBD locus and eQTLs regulating *MMRN1* expression (*MMRN1*, PPH4 = 0.86; Figure 3B; Supplementary Table 1). Sensitivity analyses confirmed that *SNCA-AS1* and *MMRN1* colocalizations were robust to changes in the prior probability of a variant associating with both traits (i.e., p_12_ prior). We did not find evidence of colocalization between the RBD *SCARB2* locus and eQTLs in brain or blood (Supplementary Figures 5-7).

As both the *SNCA-AS1* and *MMRN1* colocalizations were observed at the same RBD risk locus on chromosome 4, but in different tissues, we hypothesized that this may be due to tissue-specific regulation of *SNCA-AS1* and *MMRN1* expression. Using specificity, a measure of the proportion of a gene’s total expression attributable to one tissue or cell type, we explored the tissue- and cell-type-specific patterns of *SNCA-AS1* and *MMRN1* expression in (i) human bulk-tissue RNA-sequencing from GTEx consortium^17^ and (ii) human single-nucleus RNA-sequencing of the medial temporal gyrus from the Allen Institute for Brain Science (AIBS; 7 cell types).^18^ At the tissue level, *SNCA-AS1* expression was predominantly brain-specific, while *MMRN1* expression was most specific to thyroid, adipose and lung tissues and least specific to brain tissues (Figure 3C; Supplementary Table 2). At the cellular level, *SNCA-AS1* demonstrated neuronal specificity, while *MMRN1* was specific to microglia (Figure 3D; Supplementary Table 2). Together, these findings support the hypothesis that *SNCA-AS1* and *MMRN1* expression is regulated differently across cell types and tissues, and in turn, suggest that the RBD risk locus at chromosome 4 may operate differently depending on the cell type or tissue investigated.

### Polygenic risk scores distinguish iRBD and PD with RBD from PD without RBD

Next, we examined whether RBD-specific PRS can distinctly identify RBD as opposed to PD without RBD. We withheld a testing set of iRBD cases (N=212) and controls (N=1,265), re-performed the GWAS and meta-analysis with the remaining samples, then created a polygenic risk profile for RBD using all independent variants (R2>0.1) which passed GWAS FDR correction (p<1E-05, 47 variants, Supplementary Table 3). We calculated PRS in the withheld testing set of iRBD cases and controls, as well as independent cohorts of PD+pRBD and PD without pRBD (PD-pRBD) patients and controls. In iRBD, the PRS can differentiate between iRBD cases and controls with an area under the curve (AUC) of 0.61 (95% CI 0.58-0.65, Figure 1B), on par with recent PD PRS performance.^19^ In PD+pRBD, predictive performance was similar to iRBD with an AUC of 0.60 (95% CI 0.56-0.63), compared to decreased predictive power in PD-pRBD with an AUC=0.55 (95% CI=0.51-0.59, Figure 1B).

We then divided the RBD PRS into quartiles in the iRBD, PD+pRBD, and PD-pRBD cohorts, and performed logistic regression against the phenotype (Figure 1C). In iRBD, those in the top quartile for RBD PRS were 2.9 times more likely to have RBD than those in the bottom quartile (95% CI 1.87-4.66, p=3.5E-06), while in PD+pRBD those in the top quartile were 2.4 times more likely to have PD+pRBD (95% CI 1.62-3.56, p=1.0E-05) compared to the lowest quartile of genetic risk as a reference. This difference is likely due to the presence of both iRBD patients (i.e. individuals who had RBD prior to the onset of PD) and PD patients who developed RBD after the onset of PD, which may be genetically different. False positives of pRBD could also contribute. As noted in the AUC analysis, the RBD PRS does not distinguish well between PD-pRBD cases and controls; those in the top quartile were only 1.53 times more likely to have PD-pRBD, without statistical significance (95% CI 0.99-2.34, uncorrected p=0.053, Figure 1C). The lack of RBD PRS predictive power in PD-pRBD suggests that we are likely tagging RBD-specific loci.

We tested whether the RBD PRS is associated with changes in age at onset (AAO) or rate of conversion from RBD to overt neurodegeneration, but found no statistically significant relationship at this sample size.

### Pathway analysis reveals potential role for the autophagy-lysosomal pathway in RBD pathogenesis

To examine whether specific pathways are enriched according to the RBD GWAS results, we performed pathway enrichment analysis with WebGestalt using the GWAS-nominated genes for cellular components and biological processes (Figure 1D, Supplementary Table 4). The nominated cellular components include whole membrane (FDR corrected p=0.004), lysosome (FDR p=0.004), and vacuole (FDR p=0.005). Biological processes include positive regulation of receptor recycling (FDR p=0.037), vacuole organization (FDR p=0.037), and endocytosis (FDR p=0.039). All these nominations suggest involvement of the autophagy lysosomal pathway (ALP), a key mechanism for clearing alpha-synuclein whose disruption can lead to the alpha-synuclein aggregation seen in alpha-synucleinopathies.^20,21^

### Comparison to PD GWAS loci reveals additional loci with potential distinct effects in RBD

PD is currently the best genetically-characterized alpha-synucleinopathy due to the large sample sizes available for GWAS. We aimed to further examine how PD GWAS loci behave in RBD, given the differential effects observed in the *SNCA* and *SCARB2* loci. We compared the effect size and direction of effect of the latest PD GWAS-significant variants (N=90)^13^ to the effect sizes and direction of effect of the same variants in the summary statistics of this RBD GWAS (N=90 variants were available for comparison) and the most recent DLB GWAS^8^ (N=72).

Figure 4A-C shows the similarities and differences of effect size and direction at these loci across RBD, PD and DLB. Notably, iRBD shows marked differences at some key PD loci (Figure 4A) including *SNCA, CYLD* (encoding a deubiquitinating enzyme), and *FYN* (encoding a membrane-associated tyrosine kinase). In these loci, the direction of effect in iRBD is opposite of that seen in PD, with nominal significance in iRBD (none of these pass the GWAS significance threshold for iRBD). The PD+pRBD cohort significantly deviates from this pattern with 100% of loci showing the same direction of effect with PD (Figure 4B). However, strong PD signals, such as *SNCA* 3’ variants and *LRRK2*, are not significant in the PD+pRBD cohort despite sufficient power. Similarly, all DLB nominally significant loci share the same direction of effect with PD (Figure 4C). Yet, DLB also deviates from PD, as the *MAPT, LRRK2*, and *SNCA* 3’ loci are not statistically significant despite sufficient power. All statistics are detailed in Supplementary Table 5.

**FIGURE 4.**
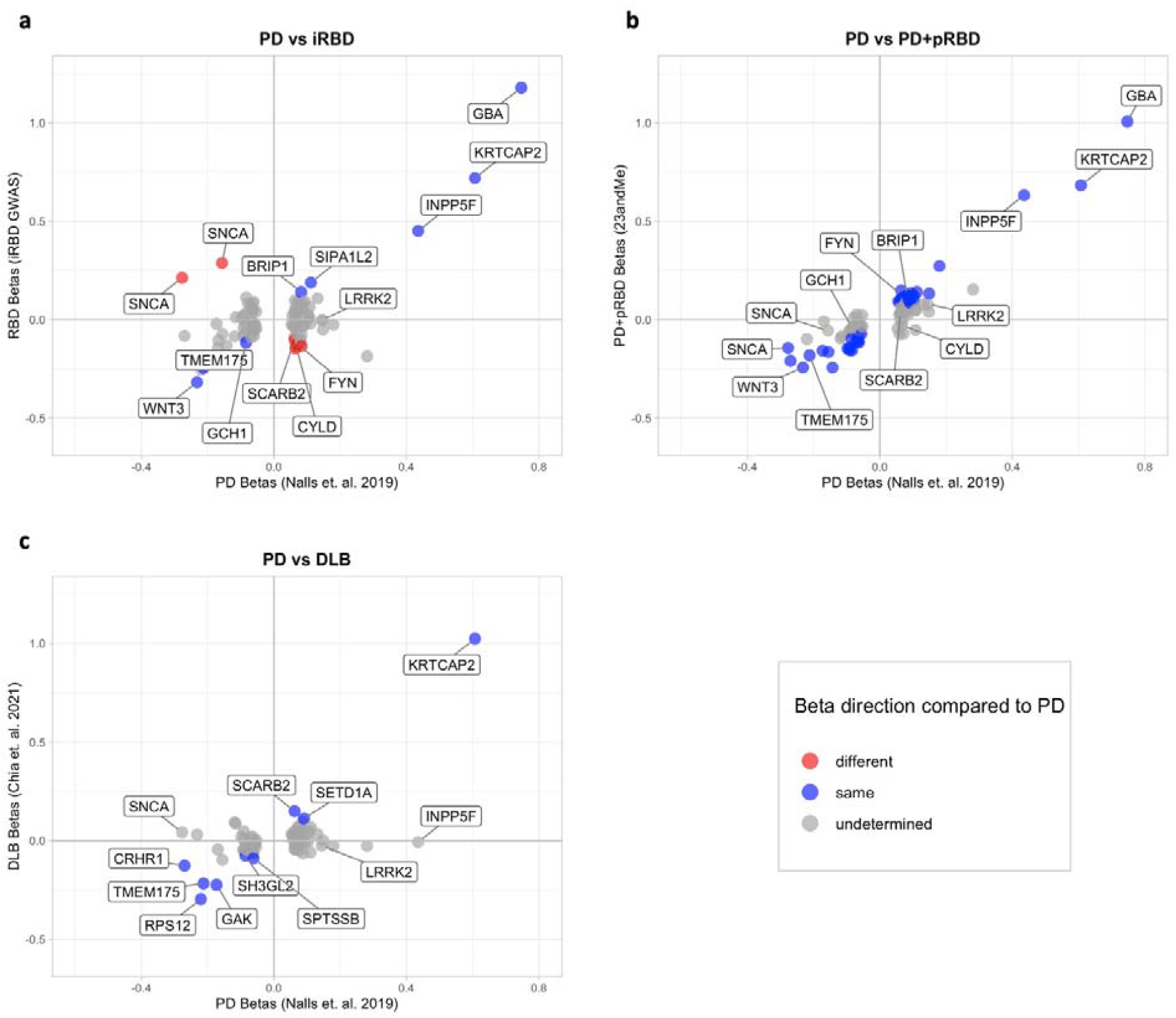
Beta-beta plots comparing synucleinopathy GWAS summary statistics to the latest PD GWAS. Colored points indicate variants with the same (blue) or opposite (red) direction of effect in both studies, with a nominally significant p-value (p<0.05). Grey points are those with undetermined direction (p>0.05 and confidence intervals cross 0). Gene names indicate the closest gene to the represented variant.

### LD score regression further supports distinct genetic background for iRBD

We used LD-score regression to examine the genetic correlation between RBD and relevant traits and exposures (Figure 5A-C). Although iRBD and PD+pRBD are positively correlated with nominal significance (rg=0.56, se=0.24, p=0.02), the two cohorts behave differently when it comes to other alpha-synucleinopathies. PD+pRBD is highly and significantly correlated with the latest PD GWAS (rg=0.76, se=0.13, p=1.2E-09). On the contrary, iRBD is not correlated with PD (rg=0.17, se=0.14, p=0.23). PD+pRBD is not correlated with DLB (rg=0.29, se=0.23, p=0.21), yet the meta-analysis of iRBD and PD+pRBD is positively correlated with DLB (rg=0.68, se=0.26, p=0.009), although it loses significance after multiple testing correction. Therefore, this possible association is likely driven by iRBD; however, LD score regression could not accurately measure genetic correlation between iRBD and DLB due to lack of sufficient power when comparing the two datasets. Additionally, iRBD is genetically correlated with Type II Diabetes (rg=0.66, se=0.23, p=0.0047), while PD+pRBD is not (rg=-0.09, se=0.10, p=0.40).

**FIGURE 5.**
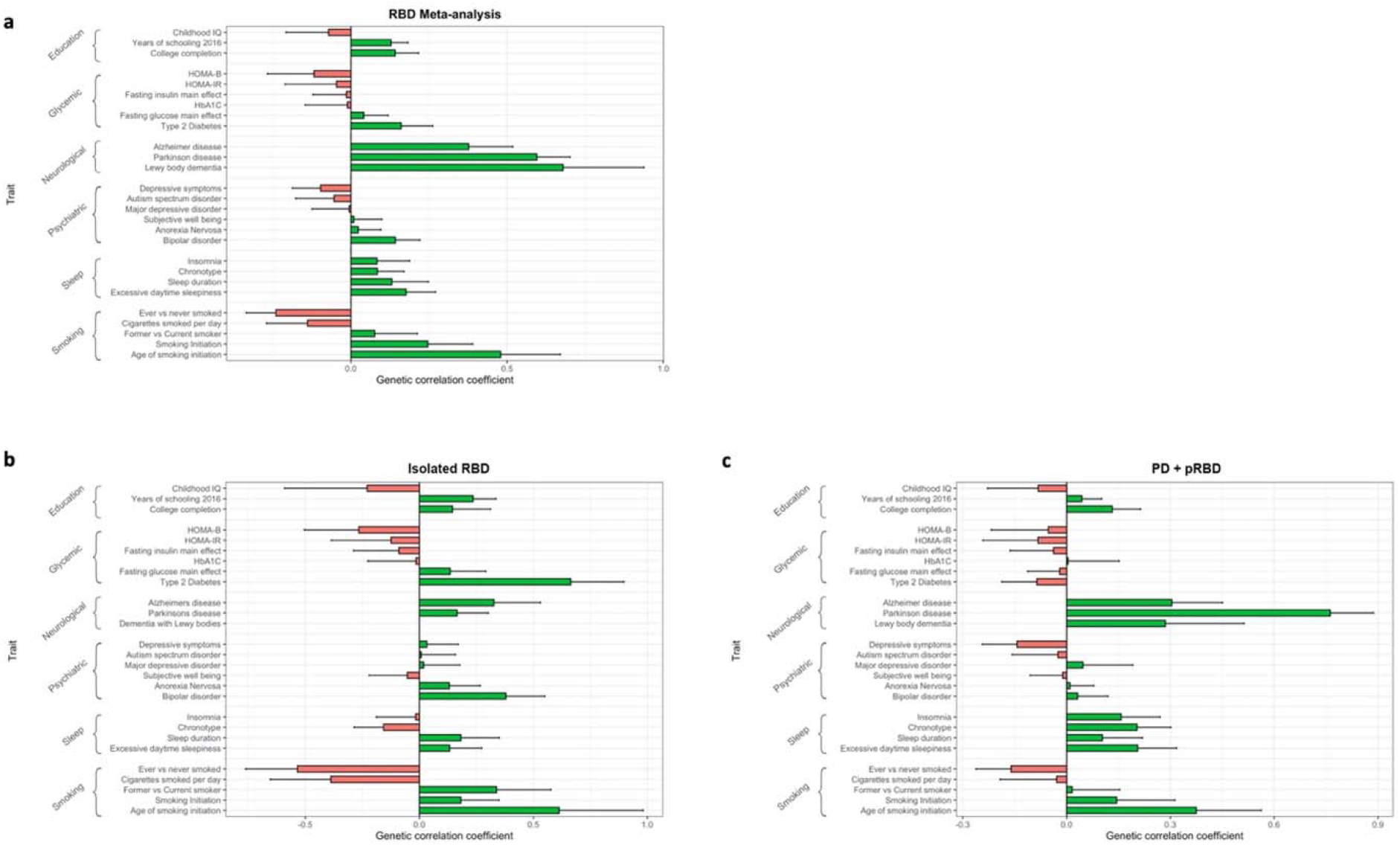
Genetic correlation results. Genetic correlation was calculated using LD-score regression for **(a)** the RBD meta-analysis, **(b)** isolated RBD (iRBD) alone, and **(c)** PD+pRBD alone. The traits tested are organized in their general categories as labeled by LD Hub. The error bars represent the standard error for the genetic correlation coefficient.

PD+pRBD and iRBD behave more similarly when discussing the following traits, so we will simply refer to the meta-analysis results. RBD shows similarities to PD with potential genetic correlations with less smoking, more education, and excessive daytime sleepiness, without significance after multiple testing correction (Supplementary Table 6). We found no genetic correlation with other sleep traits such as insomnia or sleep duration.

### Mendelian randomization suggests shared causal associations in RBD and PD

Mendelian randomization (MR) allows for the assessment of causality between an exposure and an outcome. In this case, we aimed to analyze the causal effects of traits and exposures on RBD using the RBD GWAS meta-analysis and vice-versa. Because this GWAS is not strongly powered for MR, we focused only on exposures previously linked to PD^22^ (Table 2) and variables investigated in the genetic correlation analyses (Supplementary Table 7). Similar to PD, body mass index (BMI) measurements such as arm, leg, and trunk measurements showed a potential inverse causal association (smaller measurements being potentially causal for RBD); however, significance was lost after multiple testing correction (Table 2). Other PD associations and traits from genetic correlation (e.g., Type II Diabetes and smoking) showed no causal association with RBD.

**TABLE 2.**
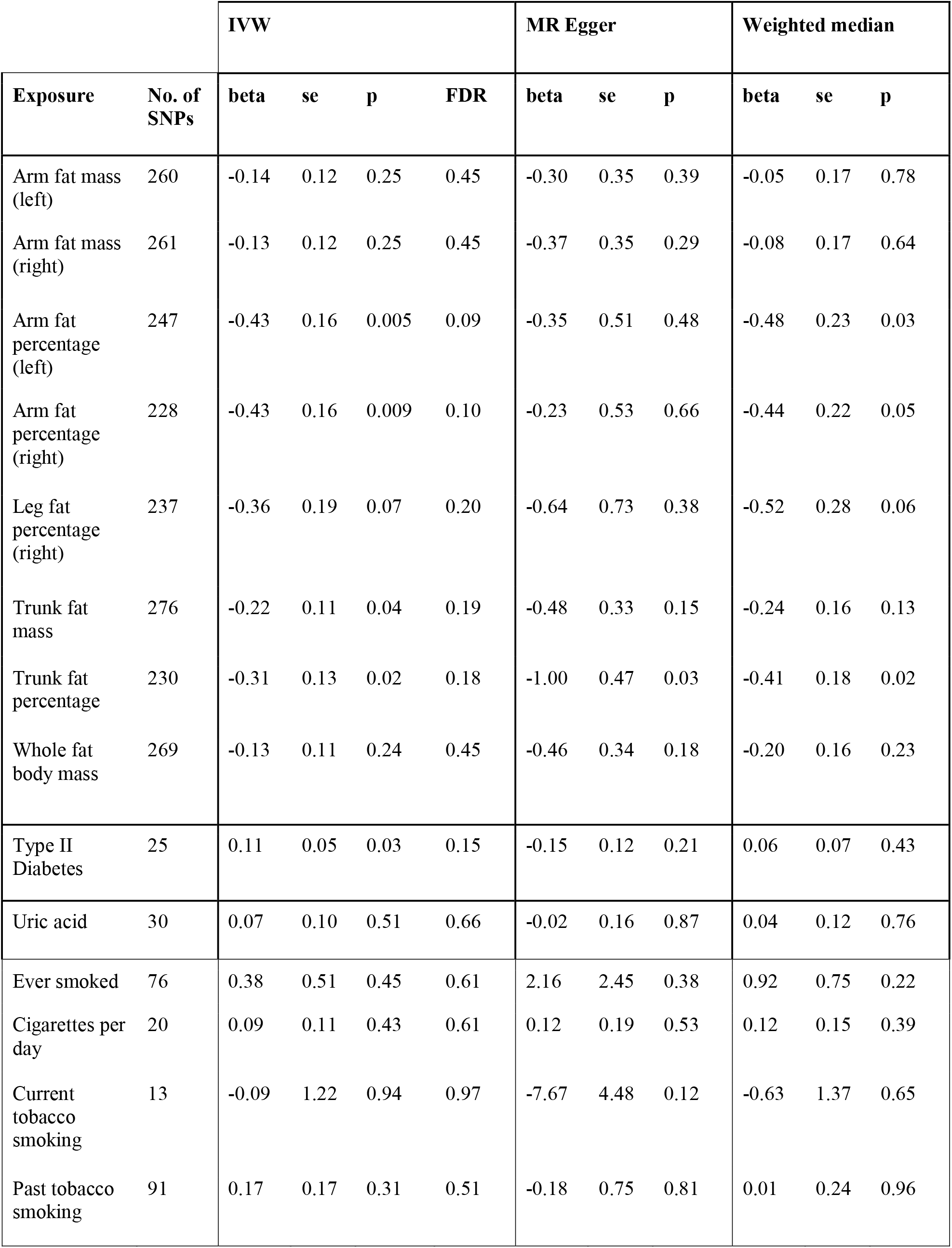
RBD MR results for traits previously linked to PD. *SNPs: single nucleotide polymorphisms; se: standard error; IVW: inverse variance weighted; MR: mendelian randomization*. Mendelian Randomization (MR) was performed to determine causal relationships between RBD and traits previously found to be associated with PD. At this sample size, no causal relationships were identified, however BMI measurements show the same direction of effect in RBD as they do in PD (lower BMI, higher risk for PD).

## DISCUSSION

In this first genome-wide study of RBD, we identified five RBD-associated loci: two novel loci near *SCARB2* and *INPP5F* and three previously reported loci near *SNCA, GBA* and *TMEM175*. Two of the loci, *SNCA* and *SCARB2*, have different and independent variants associated with RBD than those associated with PD. Colocalization and eQTL analyses suggest that these variants are associated with differential expression of *SNCA-AS1* and *SCARB2* in different brain regions, tissues, and cell types. We further show that RBD-specific PRS has different effects in individuals with iRBD or PD+pRBD compared to those with PD-pRBD. A distinct genetic background for iRBD is also supported by the lack of genetic correlation between iRBD and the most recent PD GWAS, as well as the lack of RBD association with known PD loci *LRRK2* and *MAPT*, despite sufficient power to detect them.

The differential association at the *SNCA* locus, when comparing PD and RBD, may provide a mechanistic hypothesis for gene expression-dependent regional vulnerability of different brain areas. The eQTL analysis shows that the top variant associated with RBD in the current GWAS, rs3756059, is strongly associated with reduced expression of *SNCA-AS1* in several cortical regions (Figure 2). The same risk variant has also been implicated in DLB^8^ and is in LD with a secondary PD hit in this locus which is linked to PD with dementia.^23^ When comparing PD+pRBD patients to PD-pRBD, frequency of this variant is significantly elevated in PD+pRBD.^9^ In contrast, the top variant in the recent PD GWAS, rs356182, is not associated with RBD. Additionally, we report high meta-analysis heterogeneity at this locus, which could implicate iRBD as the driver of this association. The PD+pRBD cohort, which is a mixture of PD patients who presented with RBD before PD diagnosis (iRBD) and those with symptomatic RBD later in PD progression, has a lower effect size (beta=0.15) than the iRBD cohort alone (beta=0.40). Since *SNCA-AS1* is transcribed as an antisense RNA molecule, it may lead to reduced alpha-synuclein protein expression. If this hypothesis is correct, then levels of alpha-synuclein protein may be increased in the cortical regions associated with reduced *SNCA-AS1* expression (Frontal Cortex), which could make these regions more vulnerable for neurodegeneration in carriers of the RBD-associated variant. This could explain the strong association of RBD with more rapid and more severe cognitive decline in PD,^24,25^ since the cortical brain regions associated with cognition may be more susceptible for neurodegeneration by this mechanism. Interestingly, PD patients without RBD are similar to controls when assessing cognition,^24,26^ and this variant is not a strong risk locus in this population.^9^ This hypothesized role of *SNCA-AS1* as an important, neuronally specific regulator of alpha-synuclein protein expression as a determinant of risk of alpha-synucleinopathies should be further studied.

We found a similar phenomenon in the *SCARB2* locus; rs7697073 is associated with RBD in the current GWAS, whereas in the recent PD GWAS there is an independent association at rs6825004. The PD-associated variant is possibly associated with *SCARB2* expression in the substantia nigra, while the RBD-associated variant, rs7697073, is not. This difference in *SCARB2* expression may cause an earlier degeneration of the nigrostriatal fibers in the PD cohort compared to the RBD cohort, thus explaining the earlier manifestation of motor symptoms in the former. In RBD, the top associated variant in this locus (rs7697073), like the *SNCA* locus variant, is associated more with expression in cortical brain regions, providing additional support for our hypothesis. *SCARB2*, encoding the Scavenger Receptor Class B Member 2, is the transporter of glucocerebrosidase (encoded by *GBA*) from the endoplasmic reticulum to the lysosome.^27^ It is possible that in PD this transport is affected by the variant associated with *SCARB2* expression in the substantia nigra. In RBD, this transport may be more affected in cortical regions. This could lead to specific vulnerabilities of specific brain regions in PD and RBD. For example, PD patients with RBD show significant cortical thinning when compared both to controls and to PD without RBD.^28^

Despite the inclusion of PD patients with probable RBD in the current meta-analysis, notable PD and DLB GWAS loci are absent, including *LRRK2, MAPT, BIN1*, and *APOE*. Our sample size has >80% power to detect at GWAS level significance (p<5×10-8) the associations reported in PD and DLB GWASs in these loci (supplementary figure 8). The apparent lack of association with RBD in these three important regions, which we have previously reported in candidate gene studies in smaller cohorts,^29-32^ further supports RBD as a distinct subtype, genetically and clinically. These findings suggest that PD and DLB likely include different subgroups, some of which are associated with variants in *LRRK2, MAPT* and *APOE*, while the subgroup defined by having iRBD prior to the onset of PD or DLB is not. Figure 5.6 details key overlap and distinctions between PD, DLB, and RBD GWAS loci. Additionally, we have previously shown that familial PD genes such as *PRKN, VPS35, PINK1* and others seem to have no role in iRBD,^32^ supporting distinct genetic backgrounds for iRBD and PD. The presence of subgroups within PD is also evident by the PRS analyses, showing that RBD PRS better distinguished iRBD than PD+pRBD and has minimal effect on PD-pRBD.

**FIGURE 6.**
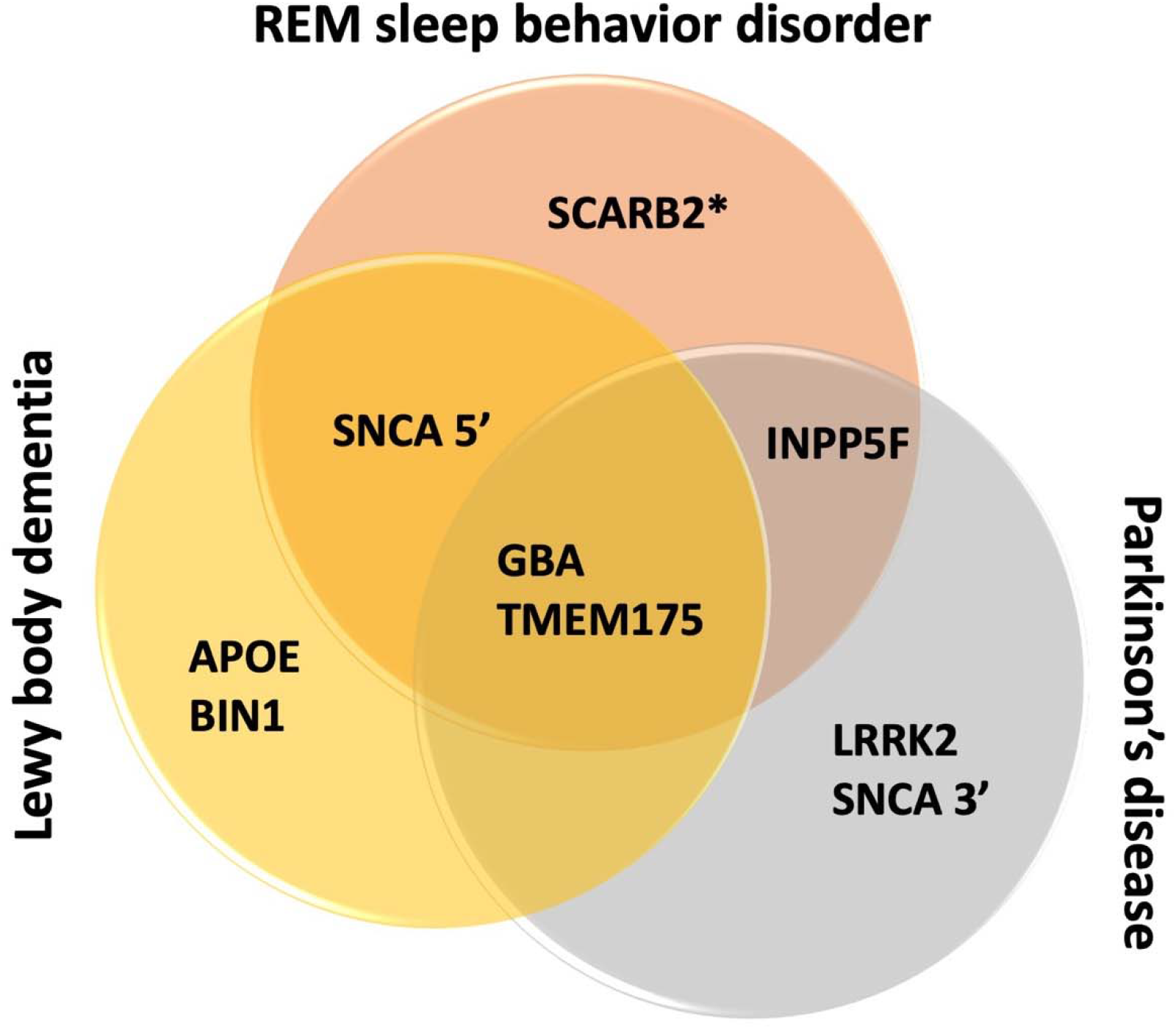
Key GWAS significant loci across three synucleinopathies. It has been shown that the genetic risk for PD and DLB do not overlap completely, and we show that the same is true for RBD and the other two synucleinopathies. Here we demonstrate key genetic risk loci for the three synucleinopathies. Only *GBA* and *TMEM175* are shared between all three, both of which play a role in the autophagy lysosomal pathway. *SNCA* plays a role in PD, DLB, and RBD risk, yet the strongest risk locus for PD is at the 3’ end of the gene while RBD and DLB share a risk locus at the 5’ end. Similarly, *SCARB2* is a risk factor for PD as well as RBD, however the RBD locus is independent of the variant identified for PD risk (as indicated by the asterisk in the figure).

Previous studies have suggested that the genetic background of iRBD may affect the rate of conversion from iRBD to overt alpha-synucleinopathies. Variants in *SNCA* and *GBA* have been associated with more rapid conversion rates.^9,10,33^ However, these results were based on a relatively small portion of the iRBD cohort with available conversion data. Therefore, in the current study we opted not to analyze individual loci due to insufficient power. When examining the effect of PRS on iRBD conversion, we did not detect a statistically significant association. Yet, it is possible that we were also underpowered, and improved PRS in future studies combined with larger numbers of iRBD patients with conversion data will yield different results.

RBD represents a distinct clinical subtype in synucleinopathies, associated with a more severe disease course in PD, DLB, and MSA, with more rapid deterioration and severe symptoms.^1,6,34-36^ Additionally, RBD is the most robust predictor of dementia in PD^25,36^ and is associated with less frequent parkinsonism at MSA onset in patients who present with RBD first.^7^ Overall, it appears that RBD is linked more closely to cognitive decline than motor symptoms in alpha-synucleinopathies. These clinical observations, combined with our genetic findings, support the presence of distinct subgroups within PD, DLB and MSA and suggest a genetic component to these distinctions. Better identification of these subgroups will be essential towards better stratification in clinical trials, and will require comprehensive genetic, biomarker, and clinical marker studies.

When considering the five loci associated with RBD in the current study, the autophagy lysosomal pathway (ALP) seems to have a major role in RBD, similar to PD and DLB. However, uniquely to RBD, four of the five nominated genes in this GWAS directly converge to the glucocerebrosidase (GBA) pathway. On top of the *GBA* locus itself, alpha-synuclein has been shown to directly interact with glucocerebrosidase products and inhibit the transport of glucocerebrosidase to the lysosome.^37,38^ Variants in the *SNCA* locus also seem to be modifiers of *GBA* penetrance and the age at onset (AAO) among *GBA* variant carriers.^39^ *SCARB2* is the transporter that carries glucocerebrosidase to the lysosome, and *TMEM175* (encoding a lysosomal potassium channel) variants affect the activity of glucocerebrosidase in humans and *in vitro*.^40^ Furthermore, *TMEM175* variants may affect the AAO of *GBA*-associated PD.^39,41^ Taken together, these previous and current findings strongly highlight the role of the glucocerebrosidase pathway in RBD. Since iRBD may appear years before the onset of overt neurodegeneration, this population is especially attractive for clinical trials aimed to prevent neurodegeneration in alpha-synucleinopathies. *GBA* targeting therapies could also be tested in this population, especially those who carry *GBA* variants.

There are several limitations to this study. First, despite being the largest RBD cohort analyzed to date, this is still a relatively small GWAS, and future RBD GWASs will likely yield additional associations. Second, the meta-analysis is PD-skewed since over half of the GWAS case cohort consists of PD+pRBD patients, some of which had iRBD prior to PD and some had PD symptoms first and RBD developed later. It is therefore possible that some signals identified by the meta-analysis are PD-enriched, and that iRBD specific signals are diluted. However, the PD+pRBD cohort notably behaves differently (e.g., the *LRRK2* and *MAPT* loci) compared to the published PD GWAS,^13^ which includes both PD with and without RBD, suggesting the subset of PD patients who have RBD may be genetically distinct. Finally, this study only includes participants of European ancestry. Future studies in other populations are vital to characterize RBD genetic risk across all ancestries, so any clinical applications can be applied universally.

In this study, we have identified novel RBD genetic loci and presented functional genomic evidence supporting regional brain vulnerability, which is dependent on differential expression of *SNCA-AS1* and *SCARB2* in different brain regions in RBD and PD. The results show that the genetic background of RBD, PD and DLB only partially overlap, and larger RBD studies will be required to better elucidate the genetic background of RBD. The present study suggests that the lysosomal pathway, and more specifically the *GBA* pathway, could be a crucial target for therapeutic development targeting RBD and aimed to prevent neurodegeneration in this population.

## METHODS

### Population

We used two cohorts for the RBD GWAS meta-analysis. The first is an iRBD cohort (N cases=1,061, N controls=8,386). The second is a cohort of PD patients with probable RBD (pRBD, N cases=1,782 and N controls=131,250), genotyped and analyzed by 23andMe. The meta-analysis combines the two for a total of 2,843 cases and 139,636 controls. We did not select individuals with pRBD but without PD from 23andMe, since a very small percentage of them will actually have iRBD; the questionnaire is not reliable for the general population but much more reliable for PD patients, albeit with false positives and false negatives. iRBD cases were collected by the International RBD Study Group and were genotyped and analyzed at McGill University. This iRBD cohort included large cohorts of French, French Canadian, Italian and British origins, and smaller cohorts from different European populations. Accordingly, for controls, we used genotype data obtained from (a) French and French-Canadian controls from McGill University (N=871); (b) the HYPERGENES Project^42^ (N=557 Italian samples); (c) the Wellcome Trust Case Control Consortium^43^ (N=5,516 British samples); (d) European control samples genotyped in the Laboratory of Neurogenetics (LNG), National Institute on Aging (NIA), National institutes of Health (NIH) (N=1,442). Principal components to adjust for population substructure were used as mentioned below. The cases were aged 68 +/-9 years (standard deviation) on average and were 81% male, and the controls were aged 58.5 +/-9 years on average, 68% male. 23andMe cases and controls were age- and sex-matched, with 83% over 60 years of age and 64.5% male. In the PRS analyses, we used an independent replication cohort of PD+/-RBD from McGill (N cases=502, N controls=907, average age 61 +/-8 years and 62% male). All cohorts used were confirmed European ancestry with principal component analysis using HapMap 3 as the reference population.

iRBD, referring to those who were diagnosed with RBD before developing overt neurodegeneration, was diagnosed according to the International Classification of Sleep Disorders (2^nd^ or 3^rd^ Edition), including video polysomnography. In the PD cohorts (except the 23andMe cohort, see below), PD was diagnosed by movement disorder specialists in accordance with the UK Brain Bank Criteria^44^ or International Parkinson Disease and Movement Disorders Society^45^ criteria. Within the PD cohorts, pRBD was identified using either the RBD single-question screen (RBD1Q)^46^ or the RBD screening questionnaire (RBDSQ),^47^ both with high sensitivity and specificity in PD.^48^ All study participants signed informed consent forms, and the study protocol was approved by the institutional review boards.

### Power calculation

To examine the power of our cohort to detect associations at GWAS significance level (p<5E-8), we used the Genetic Association Study (GAS) Power Calculator (http://csg.sph.umich.edu/abecasis/cats/gas_power_calculator/)^49^ with the following parameters: patients n=2,800, controls n=100,000, disease prevalence 0.01. We calculated the power to detect association of variants in all frequencies, with Ors of 1.2, 1.25, 1.3, 1.4 and 1.5 (Supplementary figure 8). With our sample size, we had less than 80% power to detect associations of variants at any frequency with an OR below 1.2.

### Genome-wide association study

#### 23andMe

23andMe cohorts were collected, genotyped, and filtered as previously described.^9^

#### All other cohorts

All other cohorts were prepared and analyzed as follows. The different cohorts were genotyped on the OmniExpress GWAS chips (Illumina Inc.). We performed quality control according to a standardized GWAS pipeline created at the National Institutes of Health (NIH, https://github.com/neurogenetics/GWAS-pipeline) as previously reported. To merge HYPERGENES, Wellcome Trust, and LNG controls with the McGill iRBD genotypes, we performed quality control on each cohort separately, then merged the cohorts using only variants common in all data sets. Following quality control, genotypes were filtered for minor allele frequency (MAF) > 0.01 to reduce imputation errors and imputed using Michigan Imputation Server and the Haplotype Reference Consortium^50^ r1.1 2016 reference panel (GRCh37/hg19). Only imputed genotypes with an R^2^ > 0.30 were kept for analysis, and imputed rare variants (MAF < 0.01) were excluded.

GWAS was performed using rvtests (http://zhanxw.github.io/rvtests/) single Wald association test, including sex, age, and the appropriate ancestry principal components determined by scree plot for each cohort as covariates. We implemented METAL (https://genome.sph.umich.edu/wiki/METAL_Documentation) to perform fixed-effect meta-analysis and FUMA (https://fuma.ctglab.nl/) to identify top hits according to the standard GWAS p-value threshold of p<5E-08. FUMA implemented an R^2^ threshold of 0.6 to define independent SNPs, with a subsequent R^2^ threshold of 0.1 to define lead SNPs within LD blocks. To determine whether secondary associations were present in the different loci, we used GCTA-COJO with default parameters (https://cnsgenomics.com/software/gcta/#COJO).

##### Polygenic risk score

PRSice2 (https://www.prsice.info/)^51^ and PLINK^52^ 1.9 (https://www.cog-genomics.org/plink/) were used to calculate polygenic risk scores (PRS). PRS is a summation of an individual’s genetic risk for a condition based on the effect sizes of the risk variants nominated by GWAS. To minimize overfitting, a portion of iRBD cases (N=212) and controls (N=1,265) were withheld as a testing set. The GWAS and meta-analysis were redone excluding these samples. We then determined the optimal p-value threshold for PRS in the testing set, calculated by 10,000 permutations using PRSice2, with a cautious p-value ceiling at the GWAS FDR significance threshold (p<1E-05, which was then nominated). Independent variants according to the standard pruning parameters defined by PRSice2 (R^2^>0.1) passing this threshold (N=47, Supplementary Table 3) were used to calculate the PRS. Receiver operating characteristic and area under the curve (AUC) analysis, using R version 4.0.1, was used to determine the accuracy of this PRS in differentiating between cases and controls in iRBD (the withheld samples), an independent PD+pRBD cohort (N cases=285 and N controls=902), and a PD-pRBD cohort (N cases=217 and N controls=900). Additionally, we incorporated the RBD PRS into regression models to test whether polygenic risk for RBD is associated with RBD AAO, age at diagnosis (AAD), or rate of conversion from iRBD to overt neurodegeneration as defined by both AAO and AAD. PRS code can be found on https://github.com/lynnekrohn/RBD_GWAS/blob/main/1_PRS.md.

##### Colocalization

Coloc (version 4.0.1; https://github.com/chr1swallace/coloc)^53^ was used to evaluate the probability of RBD loci and expression quantitative trait loci (eQTLs) sharing a single causal variant. Cis-eQTLs were derived from eQTLGen (accessed 19/02/2020; https://www.eqtlgen.org/cis-eqtls.html)^54^ and PsychENCODE (accessed 20/02/2020; http://resource.psychencode.org/),^55^ which represent the largest human blood and brain expression datasets, respectively (eQTLGen, N=31,684 individuals; PyschENCODE, N=1,387 individuals). For each locus, we examined all genes within 1Mb of a significant region of interest, as defined by RBD (*p*<5×10^−8^). Coloc was run using default p_1_=10^−4^ and p_2_=10^−4^ priors (p_1_ and p_2_ are the prior probability that any random SNP in the region is associated with trait 1 and 2, respectively). The p_12_ prior (the prior probability that any random SNP in the region is associated with both traits) was altered to p_12_=5×10^−6^, which has been shown to be a more robust choice than the default p_12_=10^−5^.^56^ Loci with a posterior probability of hypothesis 4 (PPH4) ≥ 0.8 were considered colocalized due to a single shared causal variant, as opposed to two distinct causal variants (PPH3). All colocalizations were subjected to sensitivity analyses using coloc’s sensitivity() function, which plots prior and posterior probabilities of each coloc hypothesis as a function of the p_12_ prior. This permits exploration of the robustness of our conclusions to changes in the p_12_ prior. Code for coloc analyses is openly available at https://github.com/RHReynolds/RBD-GWAS-analysis/.

##### Cell-type and tissue specificity measures

Specificity represents the proportion of a gene’s total expression attributable to one cell type/tissue, with a value of 0 meaning a gene is not expressed in that cell type/tissue and a value of 1 meaning that a gene is only expressed in that cell type/tissue. To determine specificity of a gene to a tissue or cell-type, specificity values from two independent gene expression datasets were generated, as described in Chia *et al*. ^8^. Briefly, these datasets included 1) bulk-tissue RNA-sequencing of 53 human tissues from the Genotype-Tissue Expression consortium (GTEx; version 8)^17^ and 2) human single-nucleus RNA-sequencing of the middle temporal gyrus from the Allen Institute for Brain Science (AIBS; https://portal.brain-map.org/atlases-and-data/rnaseq/human-mtg-smart-seq).^18^ Specificity values for GTEx were generated using modified code from a previous publication (https://github.com/jbryois/scRNA_disease),^57^ and modified to reduce redundancy among brain regions and to include protein- and non-protein-coding genes. Expression of tissues was averaged by organ (except in the case of brain). Thus, specificity values were generated for a total of 35 tissues. Specificity values for the AIBS-derived dataset were generated using gene-level exonic reads and the ‘generate.celltype.data’ function of the EWCE package (https://github.com/NathanSkene/EWCE).^58^ Specificity values for both datasets and the code used to generate these values are openly available at: https://github.com/RHReynolds/MarkerGenes.

##### Pathway Analysis

Pathway analysis was performed using functional enrichment analysis, specifically gene set enrichment, using the publicly available online tool WebGestalt (http://www.webgestalt.org/).^59^ The gene set enrichment analysis examines the enrichment of a provided set of genes in predetermined lists of genes involved in various functional pathways, detailed in supplementary table 3. An enrichment score is calculated, representing the level to which these genes are over-expressed in the various pathways, and then statistical significance is calculated using permutation testing. RBD genes included were those closest to the most significant GWAS SNP. Multiple hypothesis adjustment is applied in accordance with the false-discovery rate (FDR).

##### Heritability & Genetic Correlation

The genetic contribution to RBD, known as heritability, was calculated in clinically confirmed cases of iRBD using linkage-disequilibrium (LD) score regression on the publicly available platform LD Hub (http://ldsc.broadinstitute.org/ldhub/).^60^ Shared heritability across traits, also known as genetic correlation, was calculated using the same method and platform.^61^ Traits for shared heritability tests were chosen based on previous association to a synucleinopathy (e.g. smoking behaviors, education levels) and relevance to RBD (e.g. sleep disorders). Summary statistics for the compared traits were accessed through the LDHub platform or downloaded from publicly available sources, then formatted and analyzed using LDHub python v2.7 scripts. Bonferroni correction was calculated based on the number of traits tested (N=27, p<0.0019). LD score regression code is available on https://github.com/lynnekrohn/RBD_GWAS/blob/main/4_LD-regression.md.

##### Mendelian randomization

To assess causality, we examined traits or exposures that were nominally associated with RBD through LD-score regression (before FDR correction), as well as traits previously associated with synucleinopathies, using Mendelian Randomization (MR). In brief, MR examines the combined effect of GWAS-significant genetic loci from a trait or exposure in the summary statistics of the outcome (in this case, RBD). This approach mimics a randomized control trial since genetics are randomly assigned at birth and are largely unaffected by environmental factors. We conducted MR analyses, including sensitivity tests, heterogeneity and pleiotropy assessments, and the inverse variance weighted (IVW) method, using the TwoSampleMR R package (https://mrcieu.github.io/TwoSampleMR/)^62^ in R version 4.0.1 according to protocol previously established.^22^ Summary statistics for exposures were either downloaded from the MRBase GWAS catalogue (http://www.mrbase.org/) or extracted from published reports. P values were adjusted for multiple testing using FDR. MR code is openly available on https://github.com/lynnekrohn/RBD_GWAS/blob/main/5_MR.md.

## IRB STATEMENT

Participants provided informed consent and participated in the research online, under a protocol approved by the external AAHRPP-accredited IRB, Ethical & Independent Review Services (E&I Review).

Participants were included in the analysis on the basis of consent status as checked at the time data analyses were initiated.

## DATA AVAILABILITY

Summary statistics for the iRBD data set are available for download at www.tinyurl.com/iRBDsumStats. The full GWAS summary statistics for the 23andMe discovery data set will be made available through 23andMe to qualified researchers under an agreement with 23andMe that protects the privacy of the 23andMe participants. Please visit https://research.23andme.com/collaborate/#dataset-access/ for more information and to apply to access the data.

## ACKNOWEDGEMENTS

HYPERGENES Project: https://cordis.europa.eu/project/rcn/86758_en.html

Wellcome Trust Case Control Consortium: https://www.wtccc.org.uk/

This work was financially supported by the Michael J. Fox Foundation, Parkinson’s Society Canada, the Canadian Consortium on Neurodegeneration in Aging (CCNA), and the Canada First Research Excellence Fund (CFREF) awarded to McGill University for the Healthy Brains for Healthy Lives (HBHL) program, and was supported in part by the Intramural Research Program of the NIH, National Institute on Aging (NIA), National Institutes of Health, Department of Health and Human Services; project number ZO1 AG000535, as well as the National Institute of Neurological Disorders and Stroke. The Montreal iRBD cohort is supported by the Canadian Institutes of Health Research.

We thank the members of the NINDS Neurodegenerative Diseases Research Unit and the NIA Laboratory of Neurogenetic for their collegial support and technical assistance.

We would like to thank the research participants and employees of 23andMe for making this work possible.

The following members of the 23andMe Research Team contributed to this study: Stella Aslibekyan, Adam Auton, Elizabeth Babalola, Robert K. Bell, Jessica Bielenberg, Katarzyna Bryc, Emily Bullis, Daniella Coker, Gabriel Cuellar Partida, Devika Dhamija, Sayantan Das, Sarah L. Elson, Teresa Filshtein, Kipper Fletez-Brant, Pierre Fontanillas, Will Freyman, Pooja M. Gandhi, Karl Heilbron, Barry Hicks, David A. Hinds, Ethan M. Jewett, Yunxuan Jiang, Katelyn Kukar, Keng-Han Lin, Maya Lowe, Jey C. McCreight, Matthew H. McIntyre, Steven J. Micheletti, Meghan E. Moreno, Joanna L. Mountain, Priyanka Nandakumar, Elizabeth S. Noblin, Jared O’Connell, Aaron A. Petrakovitz, G. David Poznik, Morgan Schumacher, Anjali J. Shastri, Janie F. Shelton, Jingchunzi Shi, Suyash Shringarpure, Vinh Tran, Joyce Y. Tung, Xin Wang, Wei Wang, Catherine H. Weldon, Peter Wilton, Alejandro Hernandez, Corinna Wong, Christophe Toukam Tchakouté.

## AUTHOR CONTRIBUTIONS

L.K., C.B., M.A.N., A.B.S., and Z.G-O. conceptualized and supervised the study. J.A.R. and K.F. performed sample preparations and validation. L.K. and K.H. performed data preparation, quality control, and genome-wide association studies, C.B. contributed to these analyses. L.K. performed meta-analysis and downstream analyses, K.S., U.R., M.A.E., E.Y., and K.B. offered data checks and support. R.H.R. performed colocalization, E.G. contributed and M.R. supervised. S.B-C. and A.N. supervised Mendelian Randomization studies. I.A., M.T.M.H., J.Y.M., J.-F.G., Y.D., G.L.G., M.V., F.J., A.B., B.H., A.S., A.I., K.S., D.K., W.O., A.J., G.P., E.A., M.F., M.P., B.H., C.T., F.S-D., V.C.C., C.C.M., A.H., L.F-S., F.D., M.V., and B.A. provided biospecimens and clinical data. S.W.S.,G.A.R., L.P., R.B.P., M.A.N., and A.B.S. provided data, equipment, and supervisory support. L.K. generated figures and wrote the initial manuscript with Z.G-O. All authors critically reviewed and edited the article.

## SUPPLEMENTARY

### Supplementary figure legends

**SUPPLEMENTARY FIGURE 1. LocusZoom regional Manhattan plots for GWAS nominated RBD risk loci**. Plots were generated using LocusZoom (http://locuszoom.org/). The x-axis indicates the genomic region defined by the top hit +/-400kb, while the y axis represents the log-adjusted p values.

**SUPPLEMENTARY FIGURE 2. Sensitivity analysis of *MMRN1* colocalization**. Sensitivity analysis of colocalization between eQTLGen-derived eQTLs regulating *MMRN1* expression and RBD GWAS signals. Plot of prior (left) and posterior (right) probabilities for H0-H4 across varying p12 priors. Dashed vertical line indicates the value of p12 used in the initial analysis (p12 = 5 × 10–6), The green region in these plots show the region for which PPH4 ≥ 0.75 would still be supported.

**SUPPLEMENTARY FIGURE 3. Sensitivity analysis of *SNCA-AS1* colocalization**. Sensitivity analysis of colocalization between PscyhENCODE-derived eQTLs regulating *SNCA-AS1* expression and RBD GWAS signals. Plot of prior (left) and posterior (right) probabilities for H0-H4 across varying p12 priors. Dashed vertical line indicates the value of p12 used in the initial analysis (p12 = 5 × 10–6). The green region in these plots show the region for which PPH4 ≥ 0.75 would still be supported.

**SUPPLEMENTARY FIGURE 4. Association of RBD risk with *SNCA-AS1* expression**. Scatterplot of beta coefficients for SNPs shared between the RBD GWAS and PsychENCODE eQTLs regulating *SNCA-AS1* expression. SNPs passing genome-wide significance (p = 5 × 10–8) in the RBD GWAS and/or PyschENCODE are indicated in red. The black line represents a linear model fitted for the beta coefficients from either dataset, with the 99% confidence interval indicated with a red fill.

**SUPPLEMENTARY FIGURE 5. Regional association plot for eQTL and RBD GWAS colocalization in the region surrounding *SCARB2***. Regional association plots for eQTL (upper pane) and RBD GWAS association signals (lower pane) in the region surrounding *SCARB2*, using eQTLs derived from **(a)** the eQTLGen meta-analysis of 31,684 blood samples from 37 cohorts (PPH3 = 0.99; PPH4 = 0.01) or **(b)** PsychENCODE’s analysis of adult brain tissue from 1387 individuals (PPH3 = 0.66; PPH4 = 0.33). The x-axis denotes chromosomal position in hg19, and the y-axis indicates association p-values on a -log_10_ scale.

**SUPPLEMENTARY FIGURE 6. Sensitivity analysis of *SCARB2* colocalization using eQTLGen-derived eQTLs**. Sensitivity analysis of colocalization between eQTLGen-derived eQTLs regulating *SCARB2* expression and RBD GWAS signals. Plot of prior (left) and posterior (right) probabilities for H0-H4 across varying p_12_ priors. Dashed vertical line indicates the value of p12 used in the initial analysis (p12 = 5 × 10–6). The green region in these plots show the region for which PPH4 ≥ 0.75 would still be supported.

**SUPPLEMENTARY FIGURE 7. Sensitivity analysis of *SCARB2* colocalization using PsychENCODE-derived eQTLs**. Sensitivity analysis of colocalization between PsychENCODE-derived eQTLs regulating *SCARB2* expression and RBD GWAS signals. Plot of prior (left) and posterior (right) probabilities for H0-H4 across varying p_12_ priors. Dashed vertical line indicates the value of p12 used in the initial analysis (p12 = 5 × 10–6). The green region in these plots show the region for which PPH4 ≥ 0.75 would still be supported.

**SUPPLEMENTARY FIGURE 8. Power calculations by effect size and allele frequency**. The figure shows the power of our cohort to detect associations at GWAS significance level (p<5E-8) according to the Genetic Association Study (GAS) Power Calculator.

### Supplementary tables

**SUPPLEMENTARY TABLE 1**. Results of colocalization analysis using the RBD GWAS and eQTLs derived from eQTLGen and PsychENCODE.

**SUPPLEMENTARY TABLE 2**. Specificity values of *MMRN1* and *SNCA-AS1* in GTEx and AIBS datasets.

**SUPPLEMENTARY TABLE 3**. Polygenic risk profile for RBD, comprised of 47 independents SNPs passing GWAS FDR correction (p<1E-05).

**SUPPLEMENTARY TABLE 4**. Detailed significant results from pathway analysis with WebGestalt.

**SUPPLEMENTARY TABLE 5**. GWAS summary statistics for all GWAS nominated PD loci in PD, iRBD, PD+pRBD, and DLB.

**SUPPLEMENTARY TABLE 6**. LD-score regression genetic correlation results for iRBD, PD+pRBD, and the RBD GWAS meta-analysis.

**SUPPLEMENTARY TABLE 7**. Mendelian Randomization results and sensitivity analyses for all studied traits and exposures in the RBD meta-analysis.

## REFERENCES

1. Dauvilliers, Y. et al. REM sleep behaviour disorder. Nature Reviews Disease Primers 4, 19 (2018).

2. Postuma, R. et al. Quantifying the risk of neurodegenerative disease in idiopathic REM sleep behavior disorder. 72, 1296–1300 (2009).

3. Postuma, R.B. et al. Risk and predictors of dementia and parkinsonism in idiopathic REM sleep behaviour disorder: a multicentre study. Brain 142, 744–759 (2019).

4. Högl, B., Stefani, A. & Videnovic, A. Idiopathic REM sleep behaviour disorder and neurodegeneration—an update. Nature Reviews Neurology 14, 40 (2018).

5. Vendette, M. et al. REM sleep behavior disorder predicts cognitive impairment in Parkinson disease without dementia. Neurology 69, 1843–1849 (2007).

6. Dugger, B.N. et al. Rapid eye movement sleep behavior disorder and subtypes in autopsy-confirmed dementia with Lewy bodies. 27, 72–78 (2012).

7. Giannini, G. et al. Progression and prognosis in multiple system atrophy presenting with REM behavior disorder. Neurology 94, e1828–e1834 (2020).

8. Chia, R. et al. Genome sequencing analysis identifies new loci associated with Lewy body dementia and provides insights into its genetic architecture. Nature genetics, 1–10 (2021).

9. Krohn, L. et al. Fine-mapping of SNCA in rapid eye movement sleep behavior disorder and overt Synucleinopathies. Annals of neurology 87, 584–598 (2020).

10. Krohn, L. et al. GBA variants in REM sleep behavior disorder: a multicenter study. Neurology 95, e1008–e1016 (2020).

11. Gan-Or, Z. et al. GBA mutations are associated with rapid eye movement sleep behavior disorder. Annals of clinical and translational neurology 2, 941–945 (2015).

12. Krohn, L. et al. Genetic, structural and functional evidence link TMEM175 to synucleinopathies. Annals of Neurology.

13. Nalls, M.A. et al. Identification of novel risk loci, causal insights, and heritable risk for Parkinson’s disease: a meta-analysis of genome-wide association studies. The Lancet Neurology 18, 1091–1102 (2019).

14. Consortium, G. The GTEx Consortium atlas of genetic regulatory effects across human tissues. Science 369, 1318–1330 (2020).

15. Akbarian, S. et al. The psychencode project. Nature neuroscience 18, 1707–1712 (2015).

16. van der Wijst, M.G. et al. Science Forum: The single-cell eQTLGen consortium. Elife 9, e52155 (2020).

17. Consortium, G. The Genotype-Tissue Expression (GTEx) pilot analysis: Multitissue gene regulation in humans. Science 348, 648–660 (2015).

18. Hawrylycz, M.J. et al. An anatomically comprehensive atlas of the adult human brain transcriptome. Nature 489, 391–399 (2012).

19. Nalls, M.A. et al. Large-scale meta-analysis of genome-wide association data identifies six new risk loci for Parkinson’s disease. Nature genetics 46, 989 (2014).

20. Martini-Stoica, H., Xu, Y., Ballabio, A. & Zheng, H. The autophagy–lysosomal pathway in neurodegeneration: a TFEB perspective. Trends in neurosciences 39, 221–234 (2016).

21. Gan-Or, Z., Dion, P.A. & Rouleau, G.A. Genetic perspective on the role of the autophagy-lysosome pathway in Parkinson disease. Autophagy 11, 1443–1457 (2015).

22. Noyce, A.J. et al. The Parkinson’s Disease Mendelian randomization research portal. Movement Disorders 34, 1864–1872 (2019).

23. Guella, I. et al. α-synuclein genetic variability: A biomarker for dementia in Parkinson disease. Annals of neurology 79, 991–999 (2016).

24. Jozwiak, N. et al. REM sleep behavior disorder and cognitive impairment in Parkinson’s disease. Sleep 40(2017).

25. Anang, J.B. et al. Predictors of dementia in Parkinson disease: a prospective cohort study. Neurology 83, 1253–1260 (2014).

26. Gagnon, J.F. et al. Mild cognitive impairment in rapid eye movement sleep behavior disorder and Parkinson’s disease. Annals of Neurology: Official Journal of the American Neurological Association and the Child Neurology Society 66, 39–47 (2009).

27. Gonzalez, A., Valeiras, M., Sidransky, E. & Tayebi, N. Lysosomal integral membrane protein-2: a new player in lysosome-related pathology. Molecular genetics and metabolism 111, 84–91 (2014).

28. Rahayel, S. et al. Brain atrophy in Parkinson’s disease with polysomnography-confirmed REM sleep behavior disorder. Sleep 42, zsz062 (2019).

29. Li, J. et al. Full sequencing and haplotype analysis of MAPT in Parkinson’s disease and rapid eye movement sleep behavior disorder. (2018).

30. Gan-Or, Z. et al. The dementia-associated APOE ε4 allele is not associated with rapid eye movement sleep behavior disorder. Neurobiology of aging 49, 218.e13-218. e15 (2017).

31. Fernández-Santiago, R. et al. Absence of LRRK2 mutations in a cohort of patients with idiopathic REM sleep behavior disorder. Neurology 86, 1072–1073 (2016).

32. Gan-Or, Z. et al. Parkinson’s disease genetic loci in rapid eye movement sleep behavior disorder. Journal of Molecular Neuroscience 56, 617–622 (2015).

33. Honeycutt, L. et al. Glucocerebrosidase mutations and phenoconversion of REM sleep behavior disorder to parkinsonism and dementia. Parkinsonism & related disorders 65, 230–233 (2019).

34. Jacobs, M.L. et al. Risk factor profile in Parkinson’s disease subtype with REM sleep behavior disorder. Journal of Parkinson’s disease 6, 231–237 (2016).

35. Palma, J.-A. et al. Prevalence of REM sleep behavior disorder in multiple system atrophy: a multicenter study and meta-analysis. Clinical Autonomic Research 25, 69–75 (2015).

36. Postuma, R.B. et al. Rapid eye movement sleep behavior disorder and risk of dementia in Parkinson’s disease: a prospective study. Movement disorders 27, 720–726 (2012).

37. Bae, E.-J. et al. Glucocerebrosidase depletion enhances cell-to-cell transmission of α-synuclein. Nature communications 5, 4755 (2014).

38. Do, J., McKinney, C., Sharma, P. & Sidransky, E. Glucocerebrosidase and its relevance to Parkinson disease. Molecular neurodegeneration 14, 1–16 (2019).

39. Blauwendraat, C. et al. Genetic modifiers of risk and age at onset in GBA associated Parkinson’s disease and Lewy body dementia. Brain 143, 234–248 (2020).

40. Krohn, L. et al. Genetic, structural, and functional evidence link TMEM175 to synucleinopathies. Annals of neurology 87, 139–153 (2020).

41. Blauwendraat, C. et al. Parkinson’s disease age at onset genome-wide association study: Defining heritability, genetic loci, and α-synuclein mechanisms. Movement Disorders 34, 866–875 (2019).

42. Salvi, E. et al. Genomewide association study using a high-density single nucleotide polymorphism array and case-control design identifies a novel essential hypertension susceptibility locus in the promoter region of endothelial NO synthase. Hypertension 59, 248–255 (2012).

43. Consortium, W.T.C.C. Genome-wide association study of 14,000 cases of seven common diseases and 3,000 shared controls. Nature 447, 661 (2007).

44. Hughes, A.J., Daniel, S.E., Kilford, L. & Lees, A.J. Accuracy of clinical diagnosis of idiopathic Parkinson’s disease: a clinico-pathological study of 100 cases. Journal of Neurology, Neurosurgery & Psychiatry 55, 181–184 (1992).

45. Postuma, R.B. et al. MDS clinical diagnostic criteria for Parkinson’s disease. Movement Disorders 30, 1591–1601 (2015).

46. Postuma, R.B. et al. A single-question screen for rapid eye movement sleep behavior disorder: a multicenter validation study. 27, 913–916 (2012).

47. Nomura, T., Inoue, Y., Kagimura, T., Uemura, Y. & Nakashima, K.J.S.m. Utility of the REM sleep behavior disorder screening questionnaire (RBDSQ) in Parkinson’s disease patients. 12, 711–713 (2011).

48. Skorvanek, M., Feketeova, E., Kurtis, M.M., Rusz, J. & Sonka, K. Accuracy of Rating Scales and Clinical Measures for Screening of Rapid Eye Movement Sleep Behavior Disorder and for Predicting Conversion to Parkinson’s Disease and Other Synucleinopathies. Frontiers in neurology 9(2018).

49. Johnson, J. & Abecasis, G. GAS Power Calculator: web-based power calculator for genetic association studies. bioRxiv. (Cold Spring Harbor Laboratory, 2017).

50. McCarthy, S. et al. A reference panel of 64,976 haplotypes for genotype imputation. 48, 1279 (2016).

51. Choi, S.W., Mak, T.S.-H. & O’Reilly, P.F. Tutorial: a guide to performing polygenic risk score analyses. Nature Protocols 15, 2759–2772 (2020).

52. Chang, C.C. et al. Second-generation PLINK: rising to the challenge of larger and richer datasets. 4, 7 (2015).

53. Giambartolomei, C. et al. Bayesian test for colocalisation between pairs of genetic association studies using summary statistics. PLoS Genet 10, e1004383 (2014).

54. Võsa, U. et al. Unraveling the polygenic architecture of complex traits using blood eQTL metaanalysis. BioRxiv, 447367 (2018).

55. Wang, D. et al. Comprehensive functional genomic resource and integrative model for the human brain. Science 362(2018).

56. Wallace, C. Eliciting priors and relaxing the single causal variant assumption in colocalisation analyses. PLoS genetics 16, e1008720 (2020).

57. Bryois, J. et al. Genetic identification of cell types underlying brain complex traits yields insights into the etiology of Parkinson’s disease. Nature genetics 52, 482–493 (2020).

58. Skene, N.G. & Grant, S.G. Identification of vulnerable cell types in major brain disorders using single cell transcriptomes and expression weighted cell type enrichment. Frontiers in neuroscience 10, 16 (2016).

59. Wang, J., Vasaikar, S., Shi, Z., Greer, M. & Zhang, B. WebGestalt 2017: a more comprehensive, powerful, flexible and interactive gene set enrichment analysis toolkit. Nucleic acids research 45, W130–W137 (2017).

60. Zheng, J. et al. LD Hub: a centralized database and web interface to perform LD score regression that maximizes the potential of summary level GWAS data for SNP heritability and genetic correlation analysis. Bioinformatics 33, 272–279 (2017).

61. Bulik-Sullivan, B. et al. An atlas of genetic correlations across human diseases and traits. Nature genetics 47, 1236 (2015).

62. Hemani, G. et al. The MR-Base platform supports systematic causal inference across the human phenome. elife 7, e34408 (2018).

