## Supplementary Figure for "Genome-wide association study of REM sleep behavior disorder identifies novel loci with distinct polygenic and brain expression effects"

**SUPPLEMENTARY FIGURES**

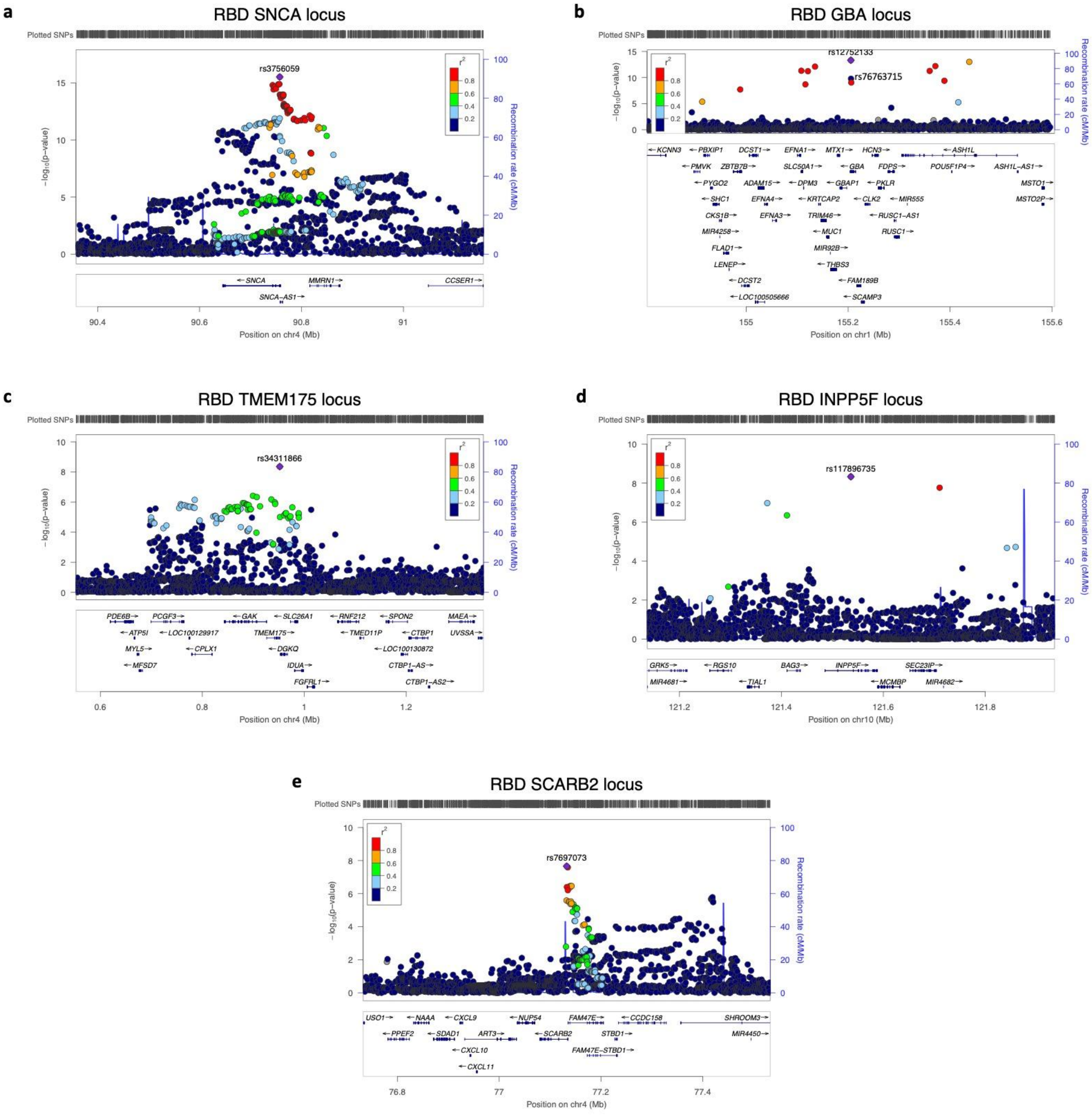


**Supplementary Figure 1. LocusZoom regional Manhattan plots for GWAS nominated RBD risk loci.** Plots were generated using LocusZoom (http://locuszoom.org/). The x-axis indicates the genomic region defined by the top hit +/- 400kb, while the y axis represents the log-adjusted p values.

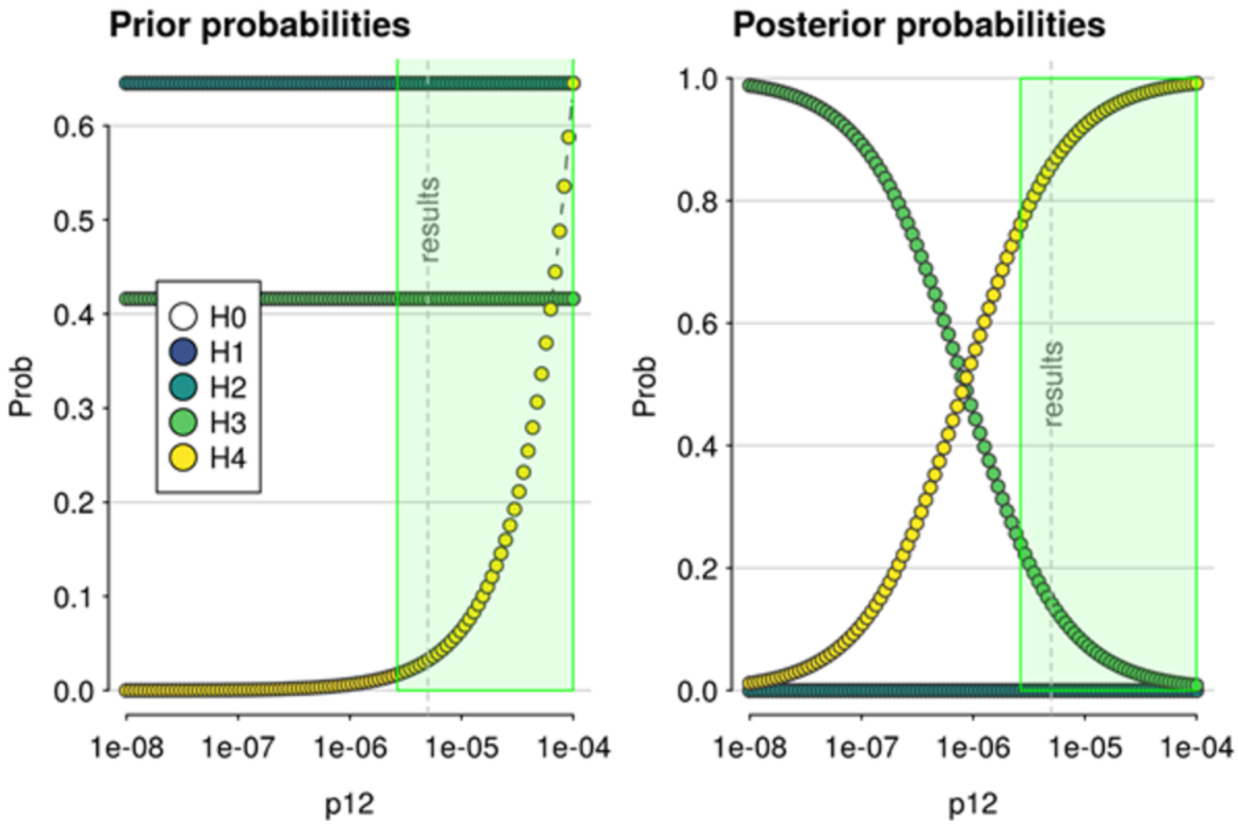


**Supplementary Figure 2. Sensitivity analysis of *MMRN1* colocalization.** Sensitivity analysis of colocalization between eQTLGen-derived eQTLs regulating MMRN1 expression and RBD GWAS signals. Plot of prior (left) and posterior (right) probabilities for H0-H4 across varying $p_{12}$ priors. Dashed vertical line indicates the value of $p_{12}$ used in the initial analysis ($p_{12} = 5 \times 10^{-6}$). The green region in these plots show the region for which PPH4 ≥ 0.75 would still be supported.

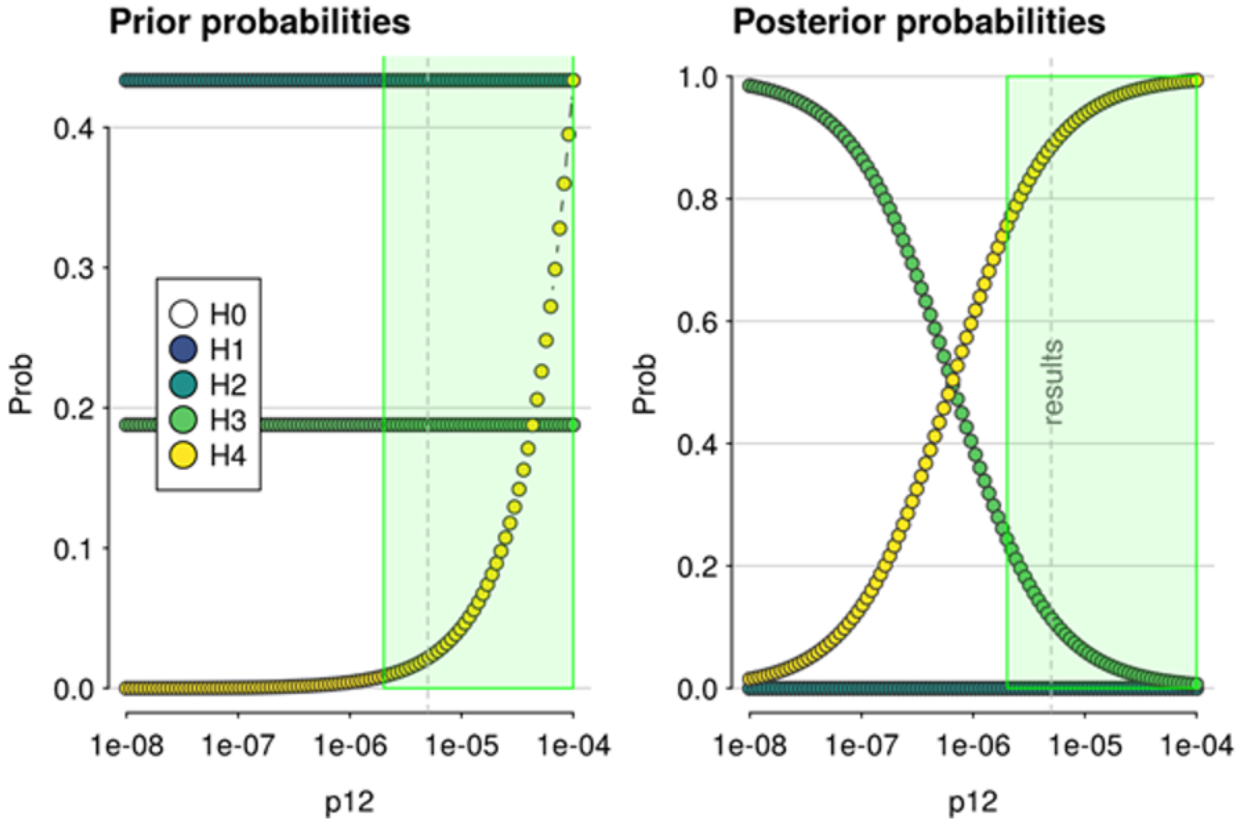


**Supplementary Figure 3. Sensitivity analysis of *SNCA-AS1* colocalization.**
Sensitivity analysis of colocalization between PscyhENCODE-derived eQTLs regulating *SNCA-AS1* expression and RBD GWAS signals. Plot of prior (left) and posterior (right) probabilities for H0-H4 across varying $p_{12}$ priors. Dashed vertical line indicates the value of $p_{12}$ used in the initial analysis ($p_{12} = 5 \times 10^{-6}$). The green region in these plots show the region for which PPH4 ≥ 0.75 would still be supported.

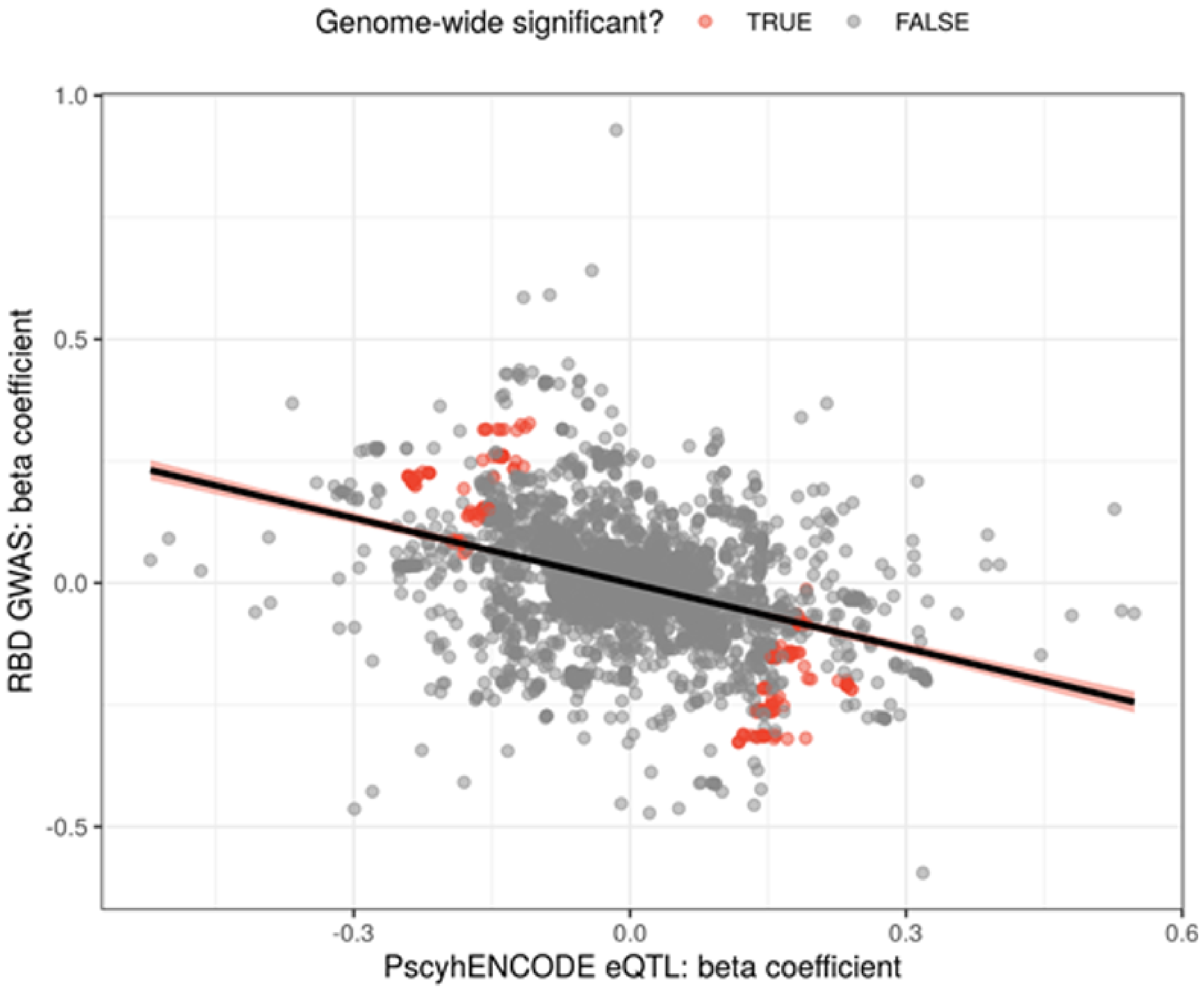


**Supplementary Figure 4. Association of RBD risk with *SNCA-AS1* expression.** Scatterplot of beta coefficients for SNPs shared between the RBD GWAS and PsychENCODE eQTLs regulating *SNCA-AS1* expression. SNPs passing genome-wide significance ($p = 5 \times 10^{-8}$) in the RBD GWAS and/or PyschENCODE are indicated in red. The black line represents a linear model fitted for the beta coefficients from either dataset, with the 99% confidence interval indicated with a red fill.

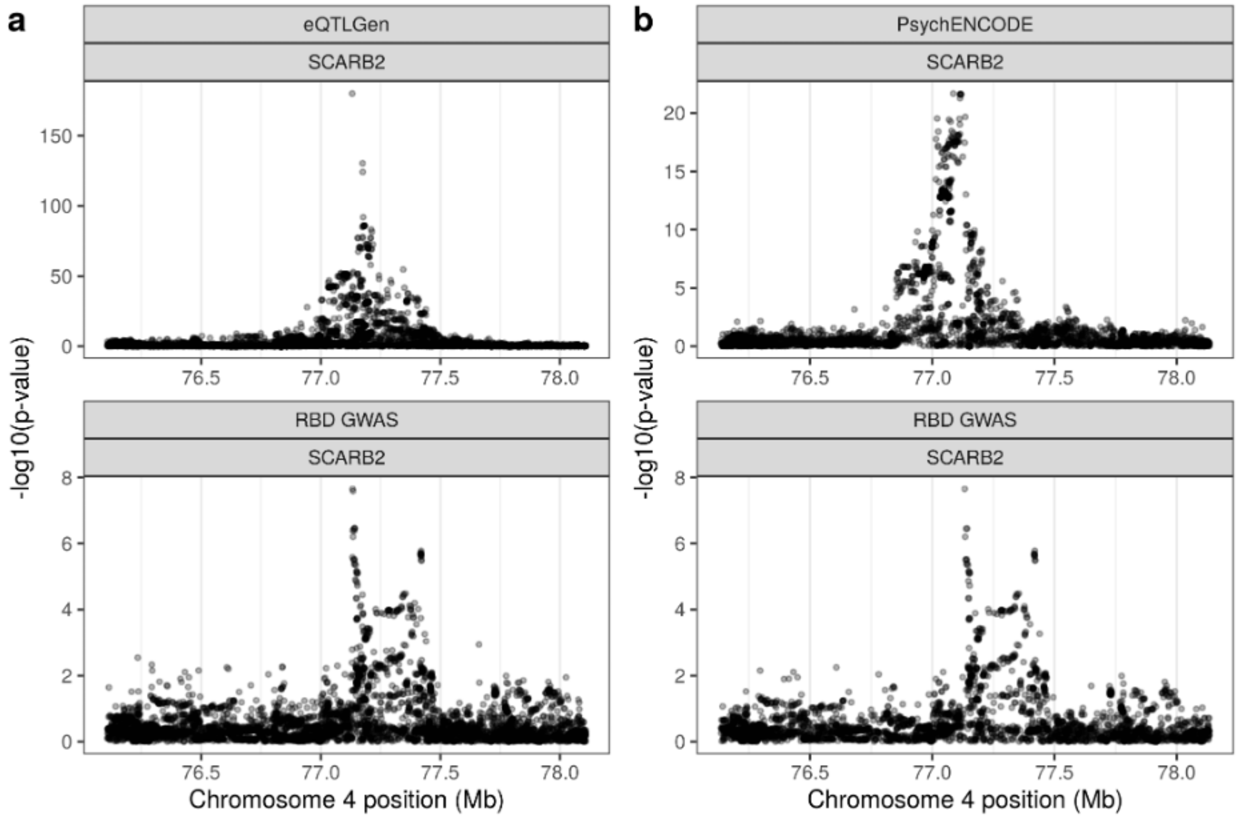


**Supplementary Figure 5. Regional association plot for eQTL and RBD GWAS colocalization in the region surrounding *SCARB2*.** Regional association plots for eQTL (upper pane) and RBD GWAS association signals (lower pane) in the region surrounding *SCARB2*, using eQTLs derived from **(a)** the eQTLGen meta-analysis of 31,684 blood samples from 37 cohorts (PPH3 = 0.99; PPH4 = 0.01) or **(b)** PsychENCODE's analysis of adult brain tissue from 1387 individuals (PPH3 = 0.66; PPH4 = 0.33). The x-axis denotes chromosomal position in hg19, and the y-axis indicates association p-values on a $-\log_{10}$ scale.

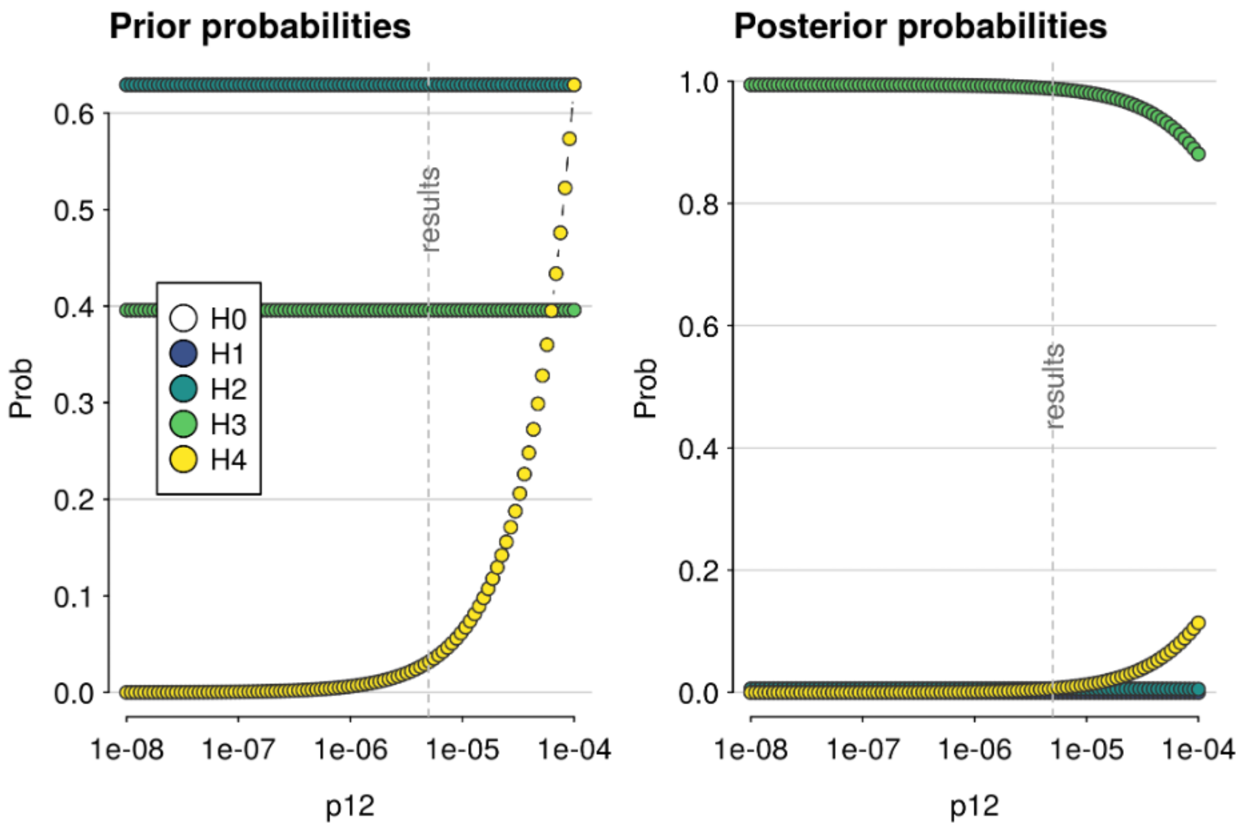


**Supplementary Figure 6. Sensitivity analysis of *SCARB2* colocalization using eQTLGen-derived eQTLs.** Sensitivity analysis of colocalization between eQTLGen-derived eQTLs regulating *SCARB2* expression and RBD GWAS signals. Plot of prior (left) and posterior (right) probabilities for H0-H4 across varying $p_{12}$ priors. Dashed vertical line indicates the value of $p_{12}$ used in the initial analysis ($p_{12}$ = 5 x $10^{-6}$). The green region in these plots show the region for which PPH4 ≥ 0.75 would still be supported.

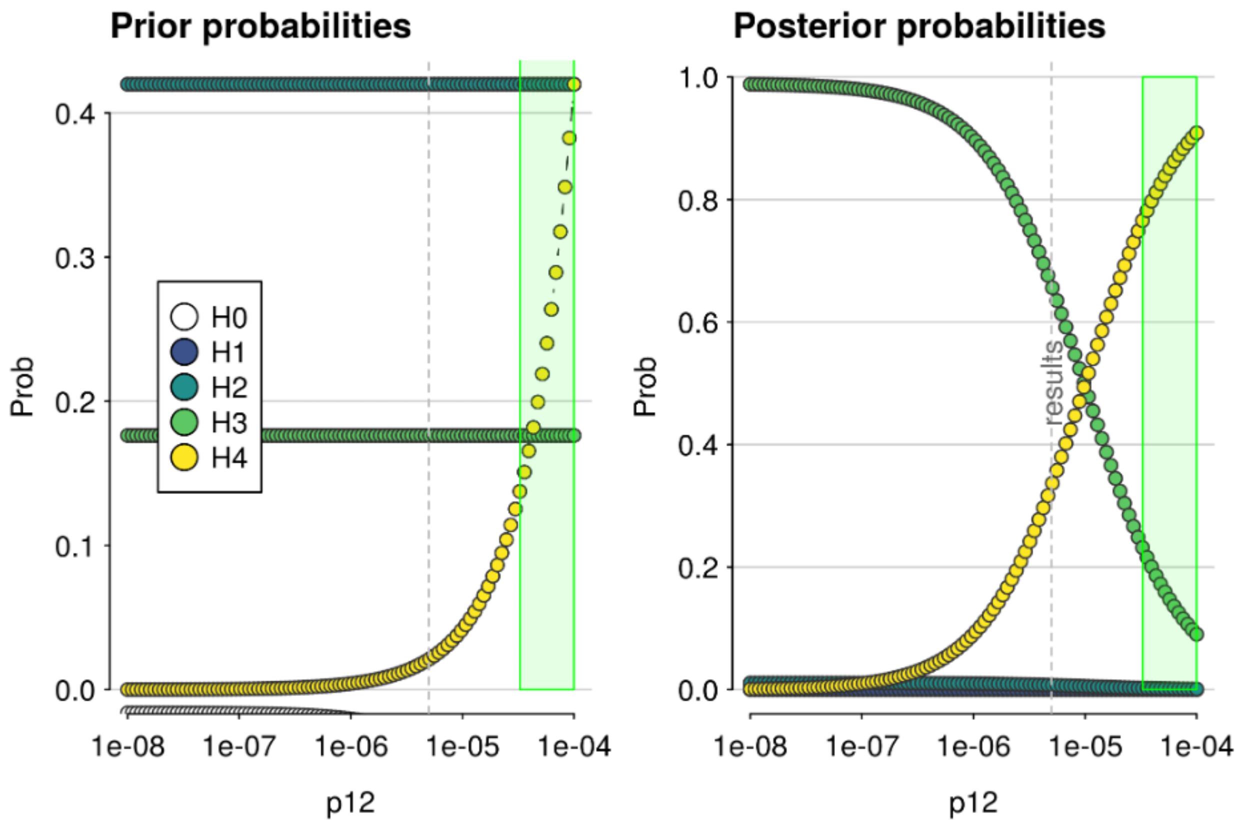


**Supplementary Figure 7. Sensitivity analysis of *SCARB2* colocalization using PsychENCODE-derived eQTLs.** Sensitivity analysis of colocalization between PsychENCODE-derived eQTLs regulating *SCARB2* expression and RBD GWAS signals. Plot of prior (left) and posterior (right) probabilities for H0-H4 across varying $p_{12}$ priors. Dashed vertical line indicates the value of $p_{12}$ used in the initial analysis ($p_{12}$ = 5 x $10^{-6}$). The green region in these plots show the region for which PPH4 ≥ 0.75 would still be supported.

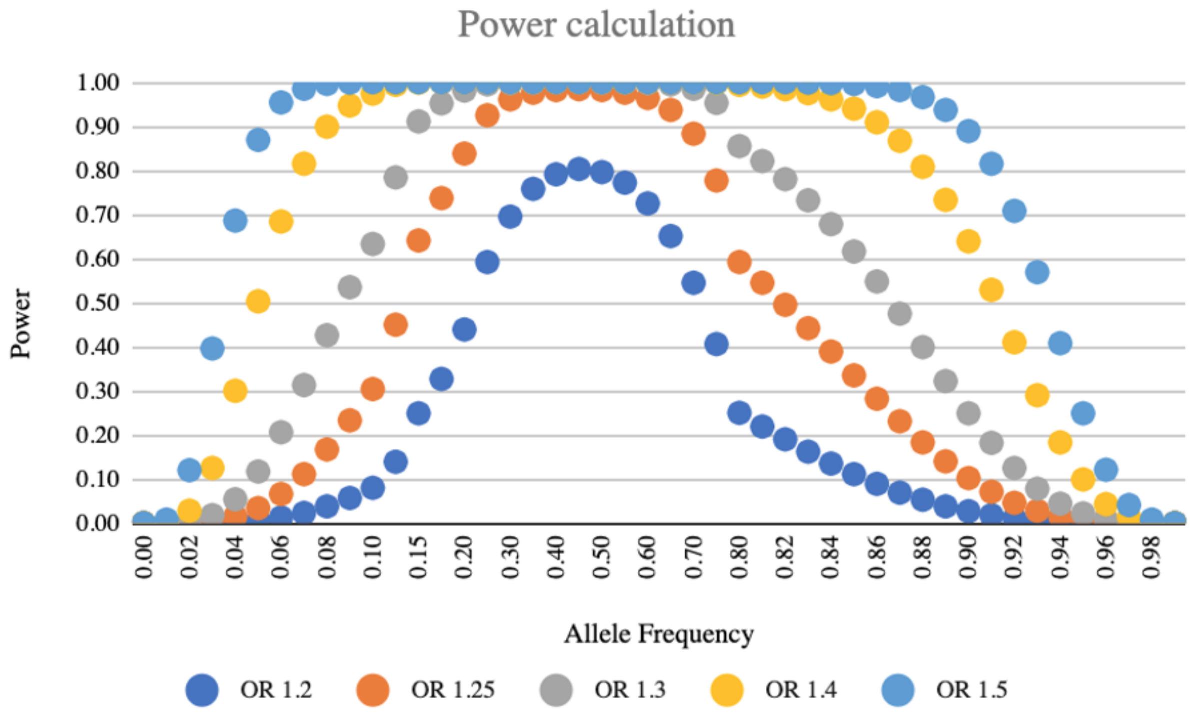


**Supplementary Figure 8. Power calculations by effect size and allele frequency.** The figure shows the power of our cohort to detect associations at GWAS significance level (p<5E-8) according to the Genetic Association Study (GAS) Power Calculator.
